# Part 1 of a 2-part, open-label clinical trial evaluating repeated nicotinamide riboside parenteral administration: 3 consecutive days of intramuscular and subcutaneous injections

**DOI:** 10.64898/2026.10.12.26365345

**Authors:** Yasmeen Nkrumah-Elie, Jun Kwon, Anne Russ, Sierra Simpson, Frehiwet Hailu, Tracy Allen, Halland Chen, Andrew Shao

**Affiliations:** ChromaDex, Inc., a Niagen Bioscience Inc. company, Los Angeles, California, USA; Impact Health Medical FL PA, Orlando, Florida, USA; Verséa Health, Inc., Tampa, Florida, USA

## Abstract

Nicotinamide riboside (NR), an endogenous precursor to the essential coenzyme nicotinamide adenine dinucleotide (NAD+), is characterized as safe and effective for longitudinally elevating NAD+ in blood and tissues, when administered orally. Interest in alternative routes of administration of NR has increased in recent years and led to the development of pharmaceutical-grade NR for intravenous and injectable administration. To evaluate the safety and efficacy of repeat daily injections of NR, an open-label, pilot, trial was initiated. Utilizing a 2×2 parallel study design, 39 study participants were randomized to intramuscular (IM) or subcutaneous (SC) injection administration, and 50 or 100 mg of NR, for 3 consecutive days, followed by a 7-day washout (50 mg IM, n=7; 100 mg IM, n=11; 50 mg SC, n=11; and 100 mg SC, n=10). To assess the primary outcome of safety, vitals, blood biomarkers, and adverse event monitoring were employed. Additional assessments included NAD+ pharmacokinetics and subjective participant-reported outcomes. Daily injections of NR were well tolerated and did not result in any unexpected adverse events or experiences. Pain and muscle soreness/tightness more than two minutes after the injection were reported by 44% and 41% of the participants, respectively, regardless of the route of administration or dose. Significant heterogeneity of pharmacokinetic parameters for NAD+ augmentation of whole blood was observed, however an average of the daily percentage increases in NAD+ was consistent across treatment groups, resulting in 9.51, 9.97, 7.83, and 9.05% for 50 mg IM, 100 mg IM, 50 mg SC, and 100 mg SC, respectively. Exploratory participant-reported outcomes suggest potential benefits on fatigue and sleep, with mixed effects on global physical health. Repeat daily injections of NR were well tolerated, and findings warrant investigation in larger, powered, placebo-controlled trials.

**Significance Statement:** Injected nicotinamide riboside (NR) was well tolerated in the study population and did not result in any treatment group safety signals. The injections increased whole blood NAD+, but the variability of timing and extent of such increase varied with each individual. The use of participant-reported outcomes suggests potential perceived benefits for fatigue, sleep, and global physical health, which require further evaluation in larger studies with a placebo control to conclude benefits in these areas.

## 1. Introduction

Nicotinamide adenine dinucleotide (NAD+) is an essential coenzyme with numerous pivotal functions in cellular metabolic processes^1^. Beyond its well-established role as a redox cofactor in cellular bioenergetics, NAD+ also serves as a critical substrate for protein families such as sirtuins and poly-ADP-ribose polymerase (PARPs), which contribute to cellular processes including DNA repair ^2^, immune signaling ^3^, and post-translational protein modification ^4,5^. Given these central roles, age-related declines in NAD+ concentrations across cells and tissues are of particular significance and may be further exacerbated by metabolic and environmental stressors such as overnutrition, excessive alcohol consumption, environmental toxicant exposure, and various disease states ^6–13^. The implications of NAD+ depletion are broad, and has been correlated with neurodegeneration, skeletal muscle dysfunction, and chronic inflammation ^9,14,15^. Mechanistically, NAD+ and related changes in tissue NAD+ flux have been associated with all of the molecular hallmarks of aging, including mitochondrial dysfunction, telomere attrition, loss of proteostasis, genomic instability, cellular senescence, stem cell exhaustion, chronic inflammation, epigenetic alterations, disabled autophagy, and dysbiosis ^9,16–29^. Therefore, raising intracellular NAD+ has attracted scientific attention as a potential therapeutic target to mitigate age-associated molecular and functional decline ^9,14,30–32^.

The provision of intravenous (IV) NAD+ as a therapeutic and health-modifying strategy was first documented by O’Hollaren in 1961^33^. Then referred to as diphosphopyridine nucleotide (DPN), O’Hollaren reported beneficial uses of DPN in individuals with substance abuse disorders ^33,34^, with subsequent investigations by an independent group of researchers exploring its application in schizophrenia^35,36^. Since then, controlled clinical trials of parenteral NAD+ have remained sparse ^37–39^. In contrast, IV NAD+’s use in wellness clinics for a variety of conditions has proliferated, largely based on anecdotal reports and clinical experience^14^. Moreover, despite its long history of clinical use, the utility of exogenously administered NAD+, whether oral or parenteral, is limited by its phosphate groups, which preclude direct cellular uptake ^40–42^. Instead, NAD+ undergoes extracellular degradation into its constituent components, such as nicotinamide (NAM) and nicotinamide riboside (NR), which subsequently enter cells through transporters present on the cellular membrane ^43,44^. Interestingly, evidence suggests acute, temporal increases in extracellular NAD+ initiate an immune response or danger signal, resulting in a proliferation of proinflammatory molecules and toxicity conferred on T-cells ^45–47^. Such a response may explain the side effects associated with IV NAD+^39,48^.

Among strategies under investigation for NAD+ repletion, the pyridine nucleoside and NAD+ precursor, NR, has emerged as a successful candidate^49–52^. A naturally occurring vitamin B3 derivative ^53^, NR undergoes direct cellular entry and feeds into the salvage NAD+ biosynthetic pathway^54^, where nicotinamide riboside kinase (NRK) catalyzes its conversion to NAD+ ^55,56^. Comparative intravenous studies have demonstrated that exogenous NR is better tolerated ^39,48^ and more effective at amplifying whole blood NAD+ than NAD+ itself ^48^. While clinical advancements in the IV administration of compounds purported to augment cellular NAD+ have continued, the use of injectable NR or NAD+ via subcutaneous (SC) or intramuscular (IM) routes remain largely unexplored, with no clinical studies in the peer-reviewed literature, to date, having systematically assessed these delivery approaches. If clinically translatable, the acute benefits of NR injections observed in preclinical models could have profound implications for the benefits of NAD+ on health outcomes ^57–59^.

Oral NR supplementation has a well-characterized safety and efficacy profile, demonstrating the ability to increase whole blood NAD+ and augment NAD+ flux in tissues, in both clinical and preclinical models ^60–66^. In healthier populations, oral NR has yielded a number of modest but promising outcomes, such as improvements in mitochondrial biogenesis and muscle myoblast differentiation ^67^, satellite cell proliferation ^67^, metabolism changes ^68^, reductions in inflammation ^63,68,69^ and substrate utilization efficiency ^68^, though several of the robust metabolic and aging benefits observed preclinically have yet to be convincingly translated to human trials ^70^. These gaps and clinical inconsistencies, alongside the complexity of the bioavailability of oral NR – driven by its partial gastrointestinal and first-pass hepatic conversion to NA and NAM ^71–73^ – have prompted interest in other delivery strategies ^57,74^. Liu et al. 2018 demonstrated that intravenous administration of NR results in greater systemic availability compared to oral delivery, leading to greater direct increases in NAD+ levels in the liver, kidney, and muscle, with secondary elevations observed in the brain^75^. Parenteral administration bypasses gastrointestinal metabolism, which may improve plasma and tissue bioavailability of NR, enabling more immediate and localized NAD+ augmentation and producing acute increases in NAD+ not observed with oral administration. Additionally, non-oral routes of NR administration may lead to more reliable, and robust increases in NAD+ at the tissue level ^75^.

A key limitation in the clinical development of parenteral NR and NAD+ is the absence of repeat-dose toxicology studies to identify target organs of toxicity and establish the no observable adverse effects level (NOAEL). A recent median lethal dose (LD50) study of parenteral NR identified route-specific effects of NR administered via IV, IM, and SC administration, with estimated LD50s of >2000 mg/kg for SC and IM routes, and 1200-2000 mg/kg for the IV route ^76^. However, a notable limitation is that the LD50 estimates only provide a reference point for acute single-dose safety, and do not substitute for repeat-dose safety studies. Accordingly, the present pilot clinical trial was conservatively designed and exploratory in nature, given in the absence of foundational sub-chronic safety data and characterization.

To evaluate the safety of Niagen® Plus (pharmaceutical-grade nicotinamide riboside chloride) as an injectable product, a two-part, open-label, randomized clinical trial was developed incorporating both daily clinical administration (Part 1) and three times per week at-home administration following a 7-day washout (Part 2, published separately). For Part 1, participants were randomized by dose and route of administration. Safety parameters including adverse events, blood biomarker, and participant experience were documented. Secondary and exploratory outcomes included NAD+ pharmacokinetics and participant-reported outcomes for fatigue, sleep, quality of life and skin. A report of the safety aspects of this study was made available in a previous preprint that combined this and a separate safety study of NR injections^77^.

## 2. Materials and Methods

### 2.1. Ethics and Regulatory Authorization

This clinical trial, “Randomized, Open-label, Safety Study of Subcutaneous and Intramuscular Injections of Niagen® Plus,” was conducted in accordance with the Declaration of Helsinki, following Good Clinical Practice (GCP) guidelines. The clinical trial was administered by Impact Health Medical FL PA (Orlando, FL) at their clinic in Aventura, Florida, USA. The study received authorization from the Institutional Review Board of the Institute of Regenerative and Cellular Medicine IRB (HHS/OHRP IRB00009500) to conduct the study under protocol number NB-2025-01 (IRB approval number: IRCM-2025-451). The trial was registered on ClinicalTrials.gov (NCT07251608). All participants provided written informed consent prior to study participation. Recruitment for the study began in September 2025 with the last participant out in February 2026. Only de-identified data was provided to the authors, outside of the clinical team, to support data analysis.

### 2.2. Trial Design

The trial was designed and conducted as a single-site, prospective, two-phase, open-label, randomized clinical trial, comparing doses and routes of administration of injectable Niagen®, clinician- and self-administered, respectively. The first phase of the study consists of four arms, 1) 50 mg NR administered IM, 2) 50 mg NR administered SC, 3) 100 mg NR administered IM, and 4) 100 mg NR administered SC. A goal of randomizing the participants 1:1:1:1 was attempted. The second phase consists of two arms, 1) 50 mg NR SC and 2) 100 mg NR SC. The purpose of this trial was to characterize differences and consistencies across NR doses and routes of administration, and to generate pilot data to inform the powering for future trials. Accordingly, statistical significance was not expected when comparing NR doses and routes of administration to baseline values (**Table 1**). Due to the exploratory, pilot nature of the trial, no formal sample size calculations were performed.

**Table 1.**
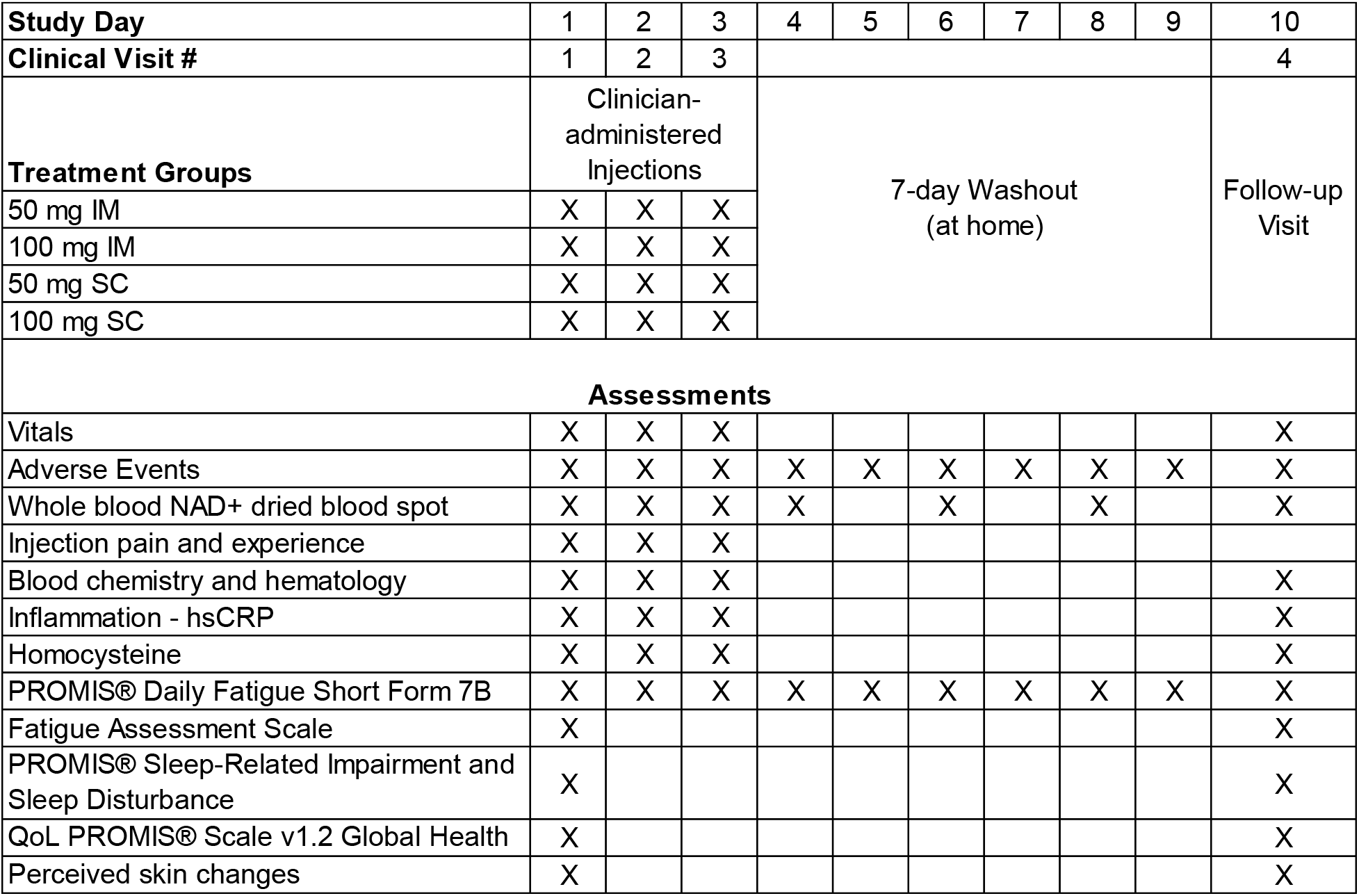
Study Design: Injection and assessment schedule.

### 2.3. Study Participants, Inclusion and Exclusion Criteria

Generally healthy adults of any gender, aged 18+, with no more than one, well-controlled, chronic condition were recruited (n=67) through IRB-approved informational documents, and community outreach. Outreach included the dissemination of digital documents at local community centers, wellness facilities, and public boards. Inclusion criteria included, 1) The demonstration of baseline fatigue as determined by an FAS score of ≥22; 2) NAD+ or NAD+ precursor naïve (including IV, IM, SC, or other administration route); 3) Non-anemic; 4) Willingness to adhere to lifestyle considerations and study procedures; 5) Ability to read English, and provide written informed consent; 6) Willingness to self-administer the study material, via subcutaneous injection for 90 days, and complete finger prick blood collections.

The participants’ lifestyle considerations required maintenance of typical diet from 2 days prior to Visit 1 until the completion of the assessments during Visit 4; maintenance of typical physical activity from 2 days prior to Visit 1 until the completion of Visit 6; avoidance of other NAD+ and NAD+ precursor products, with the exception of a multivitamin; adherence to a daily hydration requirement of ≥2L of water on the day prior to clinic visits; abstain from alcohol 24 hours prior to clinic visits; arrival at all clinic visits in a fasted state (minimum 8 hours, water only); conduct at-home assessments in a fasted state; refrain from smoking or vaping on clinic days; avoid the initiation of new medications unless medically necessary.

Exclusionary criteria included the following: 1) One or more uncontrolled chronic illnesses including diabetes, cardiovascular, liver, or kidney disease, or any form of cancer. An uncontrolled chronic illness was defined as any change to medication or other treatment modalities within the past 90 days; 2) More than one chronic disease under active treatment; 3) Any chronic disease, as determined by the primary investigator, that increases risk or confounds safety assessments; 4) Any acute illness within 14 days prior to Visit 1 (Day 1); 5) A cancer diagnosis within the past 5 years; 6) Anemia (defined as hemoglobin <100 g/L or other relevant hematologic abnormalities); 7) Current pregnancy or lactation, or unwillingness to use effective contraception if of childbearing potential; 8) Use of any NAD+ supplement, NAD+ precursor, or vitamin B_3_ product administered orally, nasally, transdermally, or by injection within the past 60 Days. NAD+ precursors and related compounds included niacin (NA), NAM, NR, nicotinamide mononucleotide (NMN), NAD+, NADH, inositol hexanicotinate, and apigenin. The exception was the use of a daily oral multivitamin that contained vitamin B_3_ (niacin/nicotinamide); 9) Hypersensitivity or allergy to NR, NA, other forms of vitamin B_3_/NAD+ precursors, or bacteriostatic water; 10) Significant aversion to needles or finger pricks; 11) Participation in another clinical intervention study within 90 Days (or 5 half-lives of the intervention, whichever is longer) prior to Visit 1 (Day 1); 12) Any other condition rendering the participant unsuitable, as determined by the investigator; 13) Excessive alcohol use, defined as 4 or more drinks on a single occasion, or illicit drug use that would prevent adherence to the protocol as determined by the investigator. Participants were not financially compensated for their participation in the study. They were provided with blood biomarkers and other results not reported in this manuscript upon study completion. In cases of clinically concerning laboratory findings, participants were notified prior to study completion and provided with their lab results.

### 2.4. Randomization and Blinding

Randomization was conducted using a computer-generated allocation schedule with concealed assignment until the time of allocation. Blinding was not incorporated into the study design or data analysis plan. Given the community-based recruitment approach, some participants were acquainted and may have discussed aspects of the study, including dosing, route of administration, and perceived outcomes, which represents a potential source of bias.

### 2.5. Investigational Product

Niagen®Plus, pharmaceutical grade nicotinamide riboside chloride, was prepared and compounded as a sterile powder for reconstitution containing 500 mg per multi-use vial by Wells Pharma (Houston, TX, USA). The study material was stored at 2–8 °C and protected from light when not in use. NR was reconstituted by the clinician staff with bacteriostatic water for the individual participants. Vials were only used for up to 28 days, according to FDA guidelines for multi-use vial products.

### 2.6. Intervention

Consented participants were instructed to drink >2 liters of water the day before clinic visits and to arrive 8-hour water-only fasted. Participants were randomized to the four-arms during the first phase, 1) 50 mg IM, 2) 50 mg SC, 3) 100 mg IM, and 4) 100 mg SC. NR administration was conducted by a qualified clinician for three consecutive days (Visits 1-3; Days 1-3). Participants were administered NR in the deltoid, upper outer quadrant for IM and in the abdomen, below the ribs and above the iliac crest, ≥5 cm from umbilicus for SC, rotating sides with each administration. The participants were subjected to a 7-day washout period and returned for visit 4 on Day 10.

### 2.7. Safety Assessments

To assess the primary outcome of the study, the safety of repeated daily NR injections included assessment of vital signs, blood biomarkers, and documentation of adverse events described by study participants and the clinical staff were collected. Vital signs (SBP, DBP, HR, RR, SpO_2_, and temp) were conducted using calibrated devices, in a standardized (seated) position. For the first NR administration, vital signs and adverse events were collected at baseline, 15-, 60-, 90-, and 180 minutes post-injection, and for visits 2 and 3, they were collected before (visit baseline) and 90-minutes post injection. Blood biomarkers were collected at baseline and 90-minutes post injection following each injection, and on Day 10, following the washout. Additionally, subjective experiences regarding tolerability and pain were collected following each injection.

### 2.8. Whole Blood NAD+ Pharmacokinetics

NR is a precursor to NAD+, thus pharmacodynamically, NR is converted intracellularly into NAD+, as its primary known function. To incorporate consistent in-clinic and at-home NAD+ testing, quantitative capillary dried blood spots (DBS) treated with a chemical stabilizer to quench enzymatic activity when blood is applied were used ^78^. Briefly, a clean and warm finger was pricked using a lancet. The first drop of blood was discarded, and the proceeding drops were collected on the DBS card. Once dried (minimum 3 hours), the card was placed in a bag with a desiccant and stored in the refrigerator or freezer. For samples collected at home, they were stored refrigerated until transport to the clinic. At the clinic, the samples were stored at −20°C until shipped, on dry ice, for LC/MS analysis by the manufacturer (Revvity Omics, Pittsburgh, PA, USA). On Day 1, dried blood spot samples were collected at baseline, 15-, 60-, 90-, and 180-minutes post injection at the clinic. On Days 2 and 3, baseline and 90-minutes post injection samples were collected at the clinic. Additional samples were collected on Days 4, 6, and 8 (at home, by participants) and on Day 10 at the clinic. Samples collected between baseline and 90 minutes were provided under fasted conditions, and every attempt was made to collect the samples at the same time, each day.

### 2.9. Participant Reported Outcomes (PRO)

#### 2.9.1. Fatigue

Fatigue was measured using two different validated subjective instruments, suitable for adult populations. The Patient-Reported Outcome Measurement Information System (PROMIS®) Short Form v1.0 – Fatigue 7b Daily measured perceived fatigue, each day throughout the study ^79,80^. Item responses were scored according to the PROMIS scoring manuals and expressed as standardized T-scores (mean = 50, SD = 10) based on the PROMIS reference population. A higher PROMIS T-score indicated greater fatigue. The 10-item Fatigue Assessment Scale (FAS) yields a total score between 10 and 50, with higher scores indicating more severe fatigue. Each item is rated on a 5-point Likert-type scale from 1 to 5. Scores are provided for two subscales, physical and mental fatigue. Total FAS scores of 22 or above are commonly cited as indicating the presence of significant fatigue ^81^. FAS was provided at screening, on Day 1 at baseline and on Day 10, Visit 4.

#### 2.9.2. Sleep

To evaluate sleep quality, participants completed the PROMIS® Sleep-Related Impairment and Sleep Disturbance questionnaire on Day 1 at baseline and on Day 10, Visit 4. This validated assessment measures both sleep impairment and sleep disturbance. Item responses were scored according to the PROMIS scoring manuals and expressed as standardized T scores (mean = 50, SD = 10) based on the PROMIS reference population^82^. A higher PROMIS T-score indicates more of the concept being measured. Therefore, higher scores indicate more sleep disturbance and sleep related impairment.

#### 2.9.3. Participant Reported Outcome: Quality of Life

The PROMIS® Global Health Scale Questionnaire was provided on Day 1 at baseline, and on Day 10, Visit 4. Domains for global mental health (GMH) and global physical health (GPH) were assessed. Item responses were scored according to the PROMIS scoring manuals and expressed as standardized T scores (mean = 50, SD = 10) based on the PROMIS reference population^83,84^. A higher PROMIS T-score indicated better perceived mental and physical health.

#### 2.9.4. Skin

A non-validated, open-ended questionnaire was provided to participants for them to assess perceived changes in both facial and whole-body skin. The following questions were provided in the questionnaire: 1) Have you observed any changes to the skin on your face since starting the injections? If yes, please describe. 2) Have others commented on any changes to the skin on your face since starting the injections? If yes, please describe. 3) Have you observed any changes to the skin on your body since starting the injections? If yes, please describe. 4) Have you noticed any other changes to your physical appearance since starting the injections? If yes, please describe.

### 2.10. Statistical Approach

The Intent-to-Treat and safety sets included all participants that received one or more injections. Statistical analysis for vitals and blood biomarkers incorporated two-group comparisons (e.g., V1 (Visit 1) Baseline vs V1 90 min): Paired t-test (scipy.stats.ttest_rel) on matched subjects. Time course comparisons involved One-way ANOVA (scipy.stats.f_oneway) for omnibus test, followed by Tukey HSD post hoc (statsmodels.stats.multicomp.pairwise_tukeyhsd) for pairwise comparisons.

Additional analyses for PRO and longitudinal assessments were developed in GraphPad Prism 11.0.2 (GraphPad Software, LLC, Boston, MA, USA), incorporating two-way ANOVA or mixed-effects analysis for repeated measures with a Tukey correction for multiple comparisons using statistical hypothesis testing for both. Due to missed participant visits and unequal sample size between the groups, a mixed-model approach was utilized to accommodate incomplete measurements. Tukey’s multiple comparisons test was used to evaluate the statistical significance within-and between-group comparisons. Statistical significance was defined as α = 0.05. Data were represented as averages ±SEM, unless otherwise noted. Statistical significance was defined as α = 0.05 was used without adjusting for multiplicity, so p-values should be read as descriptive.

## 3. Results

### 3.1. Participant Characteristics

A total of 67 individuals living in or near Aventura, Florida were recruited to participate in the study, 55 candidates were assessed for eligibility, and 40 participants were selected for randomization. One individual was excluded from receiving a dose due to hypertension at baseline. The 39 remaining participants were randomized to 4 treatment arms: 50 mg IM, 50 mg SC, 100 mg IM, or 100 mg SC. Each participant received 3 injections during the study. Clinicians administered one injection aligning with the treatment dose and route of administration each day during clinic visits 1-3 (Days 1-3). Following a 7-day washout, participants returned for Visit 4 (Day 10) for post-washout assessments (**Figure 1** and **Table 1**). All participants agreed to also participate in Part 2 of the trial, which is reported elsewhere.

**Figure 1.**
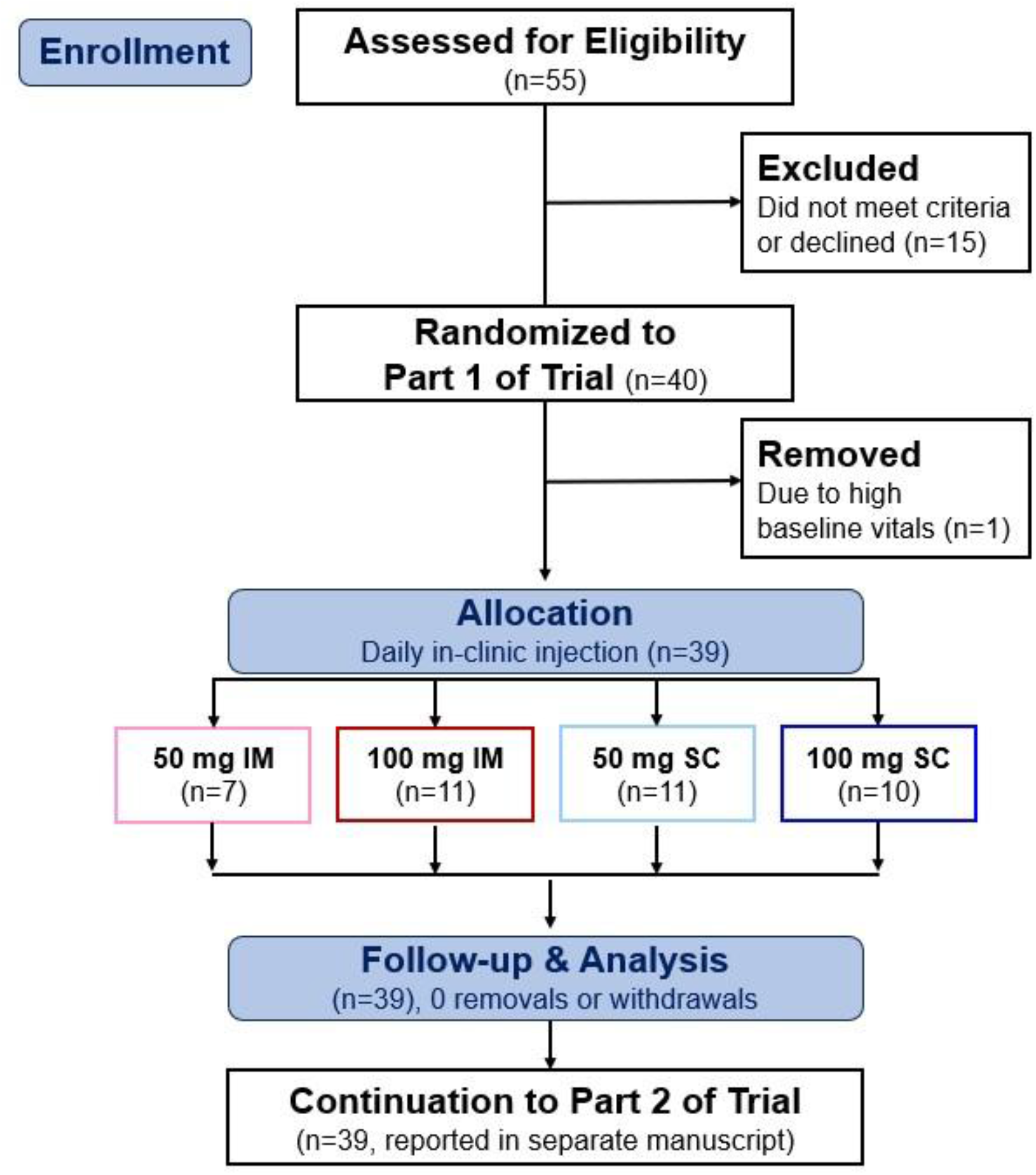
CONSORT Flow Diagram.

Female participants represented 59% of the study population, however randomization did not incorporate a gender balance, thus the 50 mg IM and 50 mg SC groups skew female, and the 100 mg IM group has more males; the 100 mg SC group is balanced for males and females. The average age of the participants was 46.7 (±1.9 SEM) and 43.6% of the participants self-reported as White, non-Hispanic. All participants had Fatigue Assessment scores ≥22, indicating some degree of more than normal fatigue as determined by the FAS. Demographics of the study population are described in **Table 2**.

**Table 2.** Baseline Participant Demographics.

| Intent to Treat (ITT), N=39 |  |  |  |  |  |
| --- | --- | --- | --- | --- | --- |
| Demographic | Injection Administration Group | 50 mg IM | 100 mg IM | 50 mg SC | 100 mg SC |
|  | n | 7 | 11 | 11 | 10 |
| Sex | Female, n (%) | 5 (71%) | 4 (36%) | 9 (82%) | 5 (50%) |
|  | Male, n (%) | 2 (29%) | 7 (64%) | 2 (18%) | 5 (50%) |
| Age | Age, years ± SEM | 44.6 ±2.65 | 47.2 ±2.74 | 46.9 ±3.95 | 47.4 ±5.22 |
| Self-Described Race/Ethnicity | Asian, n | 1 | 1 | 1 | 0 |
|  | Black & Asian | 0 | 0 | 0 | 1 |
|  | Brazilian | 0 | 1 | 0 | 0 |
|  | East Indian | 0 | 0 | 1 | 0 |
|  | Hispanic | 3 | 1 | 2 | 4 |
|  | Latin | 0 | 1 | 0 | 0 |
|  | Mixed Ethnicity (ME) | 0 | 2 | 0 | 1 |
|  | Other | 0 | 0 | 1 | 0 |
|  | White | 3 | 4 | 6 | 4 |
|  | White/ME | 0 | 1 | 0 | 0 |
| BMI, kg/m<sup>2</sup> ± SEM | | 25.1 ± 2.00 | 26.9 ± 1.48 | 24.0 ± 0.93 | 28.1 ± 2.28 |
| Smoker, n (%) |  | 0 (0%) | 1 (9.1%) | 1 (9.1%) | 1 (10%) |
| Illicit Drug Use, n (%) |  | 0 (0%) | 1 (9.1%) | 0 (0%) | 0 (0%) |
| Alcohol use | None, n (%) | 3 (42.9%) | 6 (54.5%) | 6 (54.5%) | 4 (40%) |
|  | 1-3x/wk, n (%) | 2 (28.6%) | 2 (18.2%) | 3 (27.3%) | 3 (30%) |
|  | 4-7x/wk, n (%) | 1 (14.3%) | 3 (27.3%) | 1 (9.1%) | 4 (40%) |
|  | No answer, n (%) | 1 (14.3%) | 0 (0%) | 1 (9.1%) | 0 (0%) |
| Average Screening FAS ± SEM | | 27.6 ±1.32 | 26.5 ±1.22 | 28.2 ±1.85 | 30.8 ±2.55 |

### 3.2. Vitals

Vitals were measured throughout the study to monitor participant safety, assess how the body responded to the injections, and provide as an early indicator of any adverse reactions to the injections. For the first injection vitals were measured at baseline, 15-, 60-, 90-, and 180 minutes post injection. At 90 minutes, statistically significant reductions in systolic blood pressure (SBP, ↓8.11%, p=0.011) and capillary oxygen saturation (SpO2, ↓1.68%, p=0.013) were observed in the 50 mg SC and 100 mg SC groups, respectively. At 180 minutes, a 14.67% increase in heart rate (p=0.006) was observed in the 100 mg IM group, a 5.78% decrease in respiratory rate (RR, p=0.027) in the 50 mg IM group and a 1.00% decrease (p=0.022) in SpO2 remained in the 100 mg IM group. Monitoring of vitals continued after the second and third injections, and after the washout. When comparing to Day 1 baseline and Day 10, the vitals parameters were not deemed to be statistically different (Supplement Table 1).

### 3.3. Blood Biomarkers

In addition to a standard comprehensive metabolic panel (CMP), hematology parameters for red blood cells, and platelet and white blood cell counts were included. Blood was collected for assessments on Day 1 at baseline, 90 minutes post injection on Days 1, 2, and 3, and after the washout period on Day 10. While some parameters resulted in isolated statistically significant changes (Supplement Table 2), group averages were typically within normal ranges and did not constitute a safety signal. Blood biomarkers where more than one treatment group resulted in the same statistically significant directional change at 90 minutes following the first injection were BUN and glucose. BUN decreased by 6.3 and 5.2% in the 50 mg IM and 50 mg SC groups, respectively, and glucose decreased by 7.2 and 5.2% in the 50 mg IM and 50 mg SC groups, respectively (**Figure 2A**). When comparing Day 1 to Day 10, statistically significant and directionally similar changes in blood biomarkers for multiple groups included 5.7 and 4.5% decreases in albumin, 3.0 and 3.4% decreases in hematocrit, 2.7 and 3.7% decreases in hemoglobin, and 3.0 and 2.9% decreases in red blood cells in the 50 mg SC and 100 mg IM groups, respectively (Supplement Table 2).

**Figure 2.**
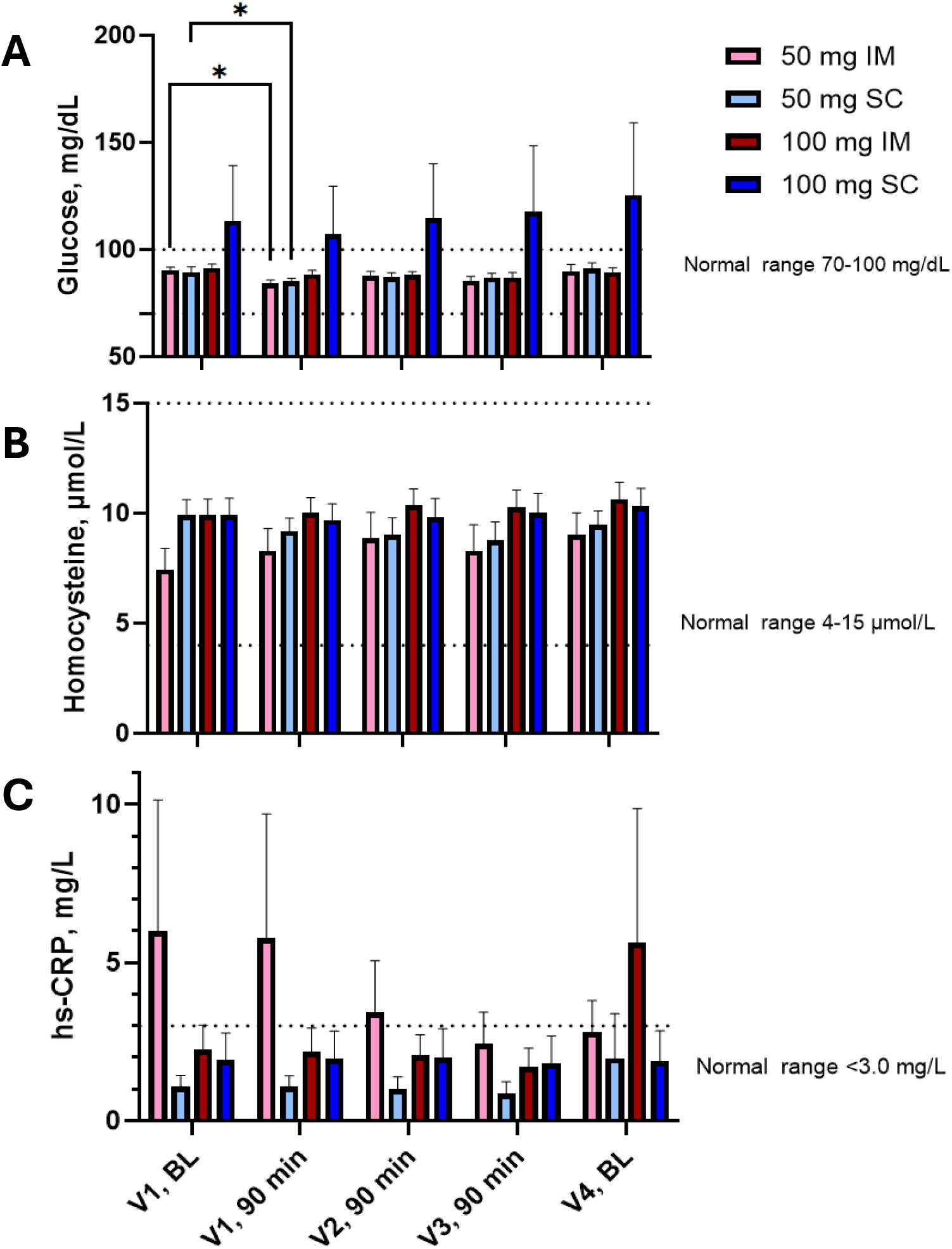
Glucose, homocysteine and hsCRP. Safety monitoring of fasted **A**) serum glucose, **B**) homocysteine, and **C**) hs-CRP were assessed at baseline on Day 1, Visit 1, as well as 90 minutes after each injection and at baseline following the 7-day washout. The normal reference ranges for each parameter are indicated as dashed lines on the figures. The data is graphed as the Average, ± SEM. Statistical analysis included a mixed effects two-way ANOVA, with Tukey’s correction for multiple comparisons. * Indicates p<0.05. *Supplementary* Figures 1-3 provide individual data demonstrating heterogeneity in responses to the injections.

Glucose, homocysteine and hsCRP were evaluated at the group and individual levels to further monitor for safety. While initial decreases in glucose were observed with the first dose, additional doses did not shift the group average throughout the study (**Figure 2**). Individually, significant heterogeneity was observed, and a clear directional response pattern could not be discerned (Supplementary Figure 1). Homocysteine levels remained within normal ranges and did not elicit clear directional changes at the group (**Figure 2B**) nor the individual (Supplementary Figure 2) level. High-sensitivity C-reactive protein (hs-CRP) was assessed to evaluate acute indications of low-level systemic inflammation. Statistically significant group changes in hs-CRP were not observed (**Figure 2C**), and in general, most participants did not exhibit significant changes in this marker. However, there were outliers in every group with hs-CRP levels that exceeded the normal range at more than one timepoint in the study, and directional pattern changes in response to NR injections were not obvious (Supplementary Figure 3).

### 3.4. Injection Experience

Participants described their injection experience immediately and 90 minutes after each injection. No unexpected adverse injection experiences were noted by the participants nor the administering clinicians. The pooled response frequency (**Figure 3**) noted “pain more than two minutes post-injection” as the most frequently described observation (44%), followed by muscle soreness/tightness (41%) and itching (31%) (**Figure 3**). A summary of participant-described experiences after each injection is provided in Supplementary **Table 3**. Redness and itching occurred following SC injections in 52.6% and 57.9% of participants, respectively. A reported increase in alertness or energy ranged from 10% in the 50 mg SC group to 71.4% for the 50 mg IM group, whereas tiredness was reported by 24.3% of all participants. Notably, the 100 mg IM cohort had 0% incidence of redness, swelling, itching, or bruising after their injection, and less than 50% incidence rate of feeling pain more than 2 minutes after injection, warmth or muscle soreness. Flushing/heat sensation was only reported with SC injections.

**Figure 3.**
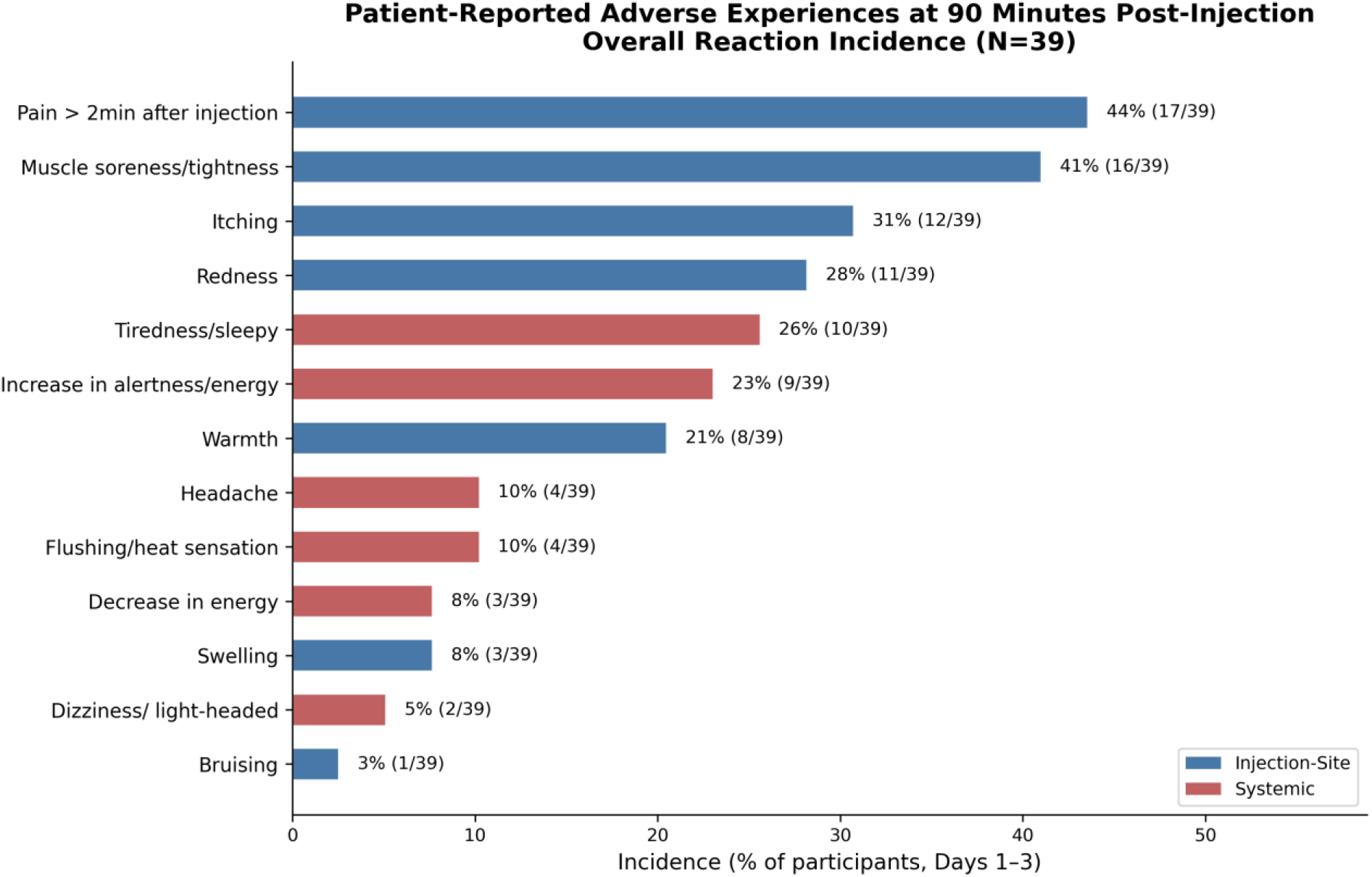
Pooled Adverse Experiences Post-Injection. Study population frequency of adverse experiences with one or more injections. Responses by treatment group are provided in *Supplementary Table 3*.

### 3.5. Adverse Events

A total of six adverse events were described; three participants developed a fever, a participant complained of a headache accompanied by slight dizziness, and a critical lab value (glucose) was reached after the washout. The severity of all adverse events was categorized as mild by the supervising medical practitioner (**Table 3**).

**Table 3.** Adverse Events.

| Route | Dose, mg | Visit | AE Description | Severity | Serious | Related to IP | Expected | Action Taken / Details |
| --- | --- | --- | --- | --- | --- | --- | --- | --- |
| SC | 050 | Visit 1 | Fever 100.6°F | Mild | No | Unknown | No | None |
| SC | 100 | Visit 1 | Headache 2/10 | Mild | No | Unknown | No | None |
| SC | 100 | Visit 1 | Slightly dizzy | Mild | No | Unknown | No | None |
| IM | 050 | Visit 2 | Temp 100.6°F at 90-min check | Mild | No | Unknown | No | Discharged |
| IM | 050 | Visit 3 | Fever after injection | Mild | No | Unknown | No | None |
| SC | 100 | Visit 4 | Critical Lab Value - Glucose | Mild | No | No | No | Participant was advised to follow-up with primary health care provider |

### 3.6. Whole Blood NAD+ Pharmacokinetics

NAD+ was measured in whole blood, using dried blood spot kits. At baseline the participants in the 50 mg IM and 50 mg SC groups had lower whole blood NAD+ levels than those in the 100 mg IM and 100 mg SC groups (**Figure 4**). Participant heterogeneity in time to maximum concentration demonstrated variability, regardless of treatment group or injection number (**Table 4**). A total of 38/39 participants demonstrated an increase in NAD+ during at least one time point assessed following the first injection, resulting in 9.5, 8.8, 11.1, and 15.1% average increases in the 50 mg IM, 50 mg SC, 100 mg IM, and 100 mg SC groups, respectively (**Figure 4**). However, when addressing observable increases in NAD+ from that day’s baseline to the observed C_max_, the average percent NAD+ increase across the three injections were 9.51, 9.97, 7.83, and 9.05% for 50 mg IM, 100 mg IM, 50 mg SC, and 100 mg SC, respectively (Figure 4D).

**Figure 4.**
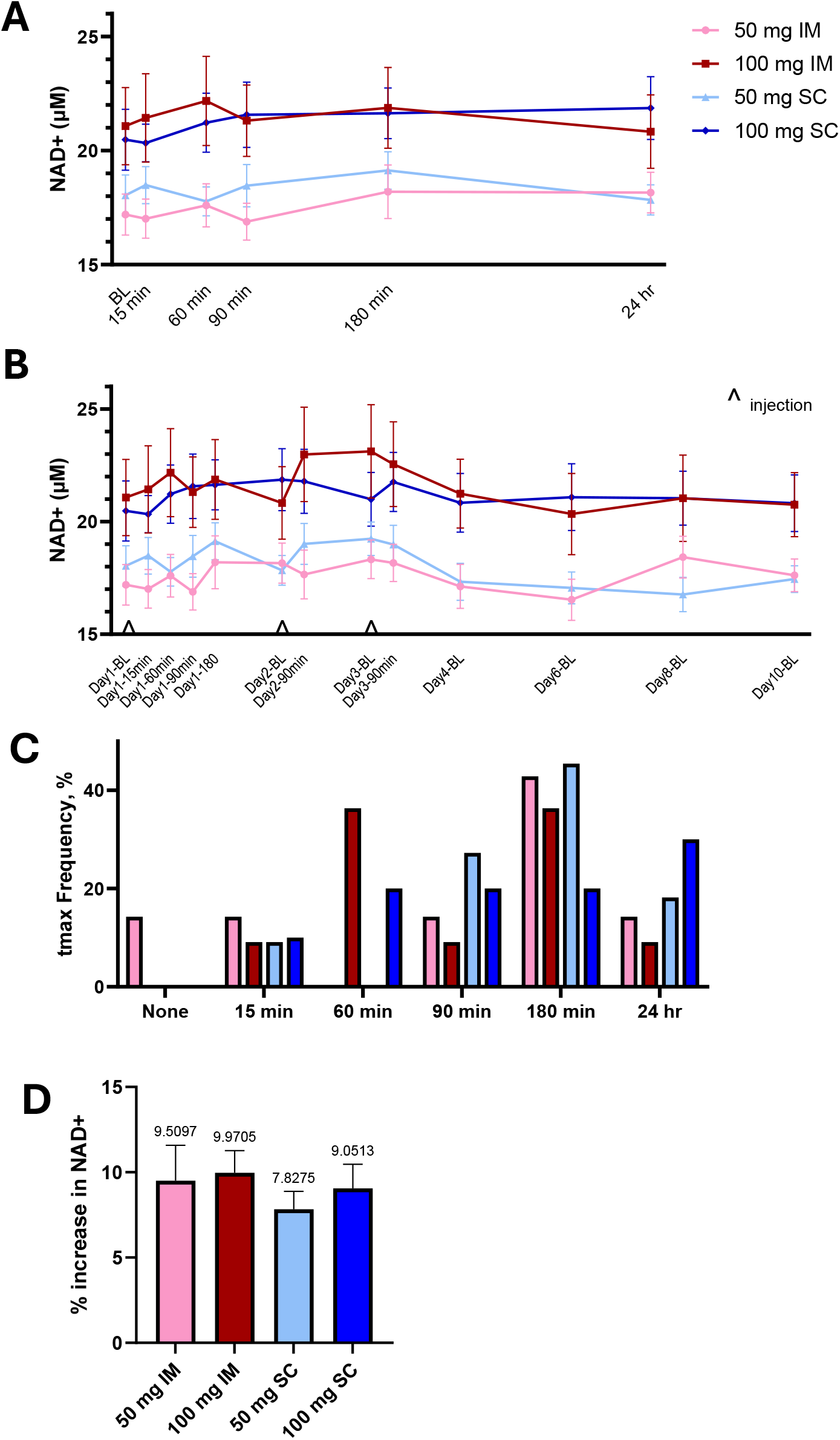
NAD+ Pharmacokinetics. Capillary whole blood NAD+ was measured by dried blood spots. **A**) Single dose NAD+ pharmacokinetics were captured at baseline (BL), 15, 60, 90, and 180 minutes and 24 hours. **B**) NAD+ variance by group for the duration of the study (visit 1-4). **C**) t_max_ frequency by participant for the first injection. **D**) Average % increase in whole blood NAD+ by treatment group for the three injections. When an increase in NAD+ concentration was not observed, a value of zero(0) was given to the C_max_. The numbers above the bars indicate the average value. Values are reported as the group average ±SEM. Statistical analysis included a mixed effects two-way ANOVA, with Tukey’s correction for multiple comparisons. * = p<0.05 and ** = p<0.01.

**Table 4.** NAD+ Pharmacokinetics: t_max_ frequency (%) for each injection by treatment group.

| Injection # | Time point | 50 mg IM | 50 mg SC | 100 mg IM | 100 mg SC |
| --- | --- | --- | --- | --- | --- |
| 1 | none | 14.29 | 0.00 | 0.00 | 0.00 |
|  | 15 min | 14.29 | 9.09 | 9.09 | 10.00 |
|  | 60 min | 0.00 | 0.00 | 36.36 | 20.00 |
|  | 90 min | 14.29 | 27.27 | 9.09 | 20.00 |
|  | 180 min | 42.86 | 45.45 | 36.36 | 20.00 |
|  | 24 hr | 14.29 | 18.18 | 9.09 | 30.00 |
| 2 | none | 28.57 | 9.09 | 9.09 | 40.00 |
|  | 90 min | 28.57 | 45.45 | 45.45 | 30.00 |
|  | 24 hr | 42.86 | 45.45 | 45.45 | 20.00 |
|  | Undetermined | 0.00 | 0.00 | 0.00 | 10.00 |
| 3 | none | 57.14 | 63.64 | 36.36 | 10.00 |
|  | 90 min | 14.29 | 18.18 | 45.45 | 30.00 |
|  | 24 hr | 28.57 | 18.18 | 18.18 | 50.00 |
|  | Undetermined | 0.00 | 0.00 | 0.00 | 10.00 |

### 3.7. Subjective Assessments of Fatigue, Sleep, Quality of Life, and Skin

Two different assessments of perceived fatigue were provided to the participants. A numerical decrease in perceived fatigue, that did not reach statistical significance, was observed throughout the study and washout period. When comparing Day 1 baseline and Day 10, a statistically significant decrease in the Fatigue T-score observed in the 100 mg IM and 100 mg SC groups. Additionally, the FAS score decreased significantly in the 100 mg SC group, suggesting improvements in fatigue. Sleep domains of sleep disturbance and sleep impairment were improved from baseline to Day 10 in the 100 mg IM and 100 mg SC groups, respectively. For the Quality-of-Life assessment, a statistically significant decrease in the domain for perceived physical health was noted in the 50 mg SC group, whereas an improvement was perceived in the 100 mg SC group. Statistically significant changes in perceived mental health were not observed during the study (**Figure 5**). Participants did not notice changes in their facial or body skin quality from baseline to Day 10 (data not shown).

**Figure 5.**
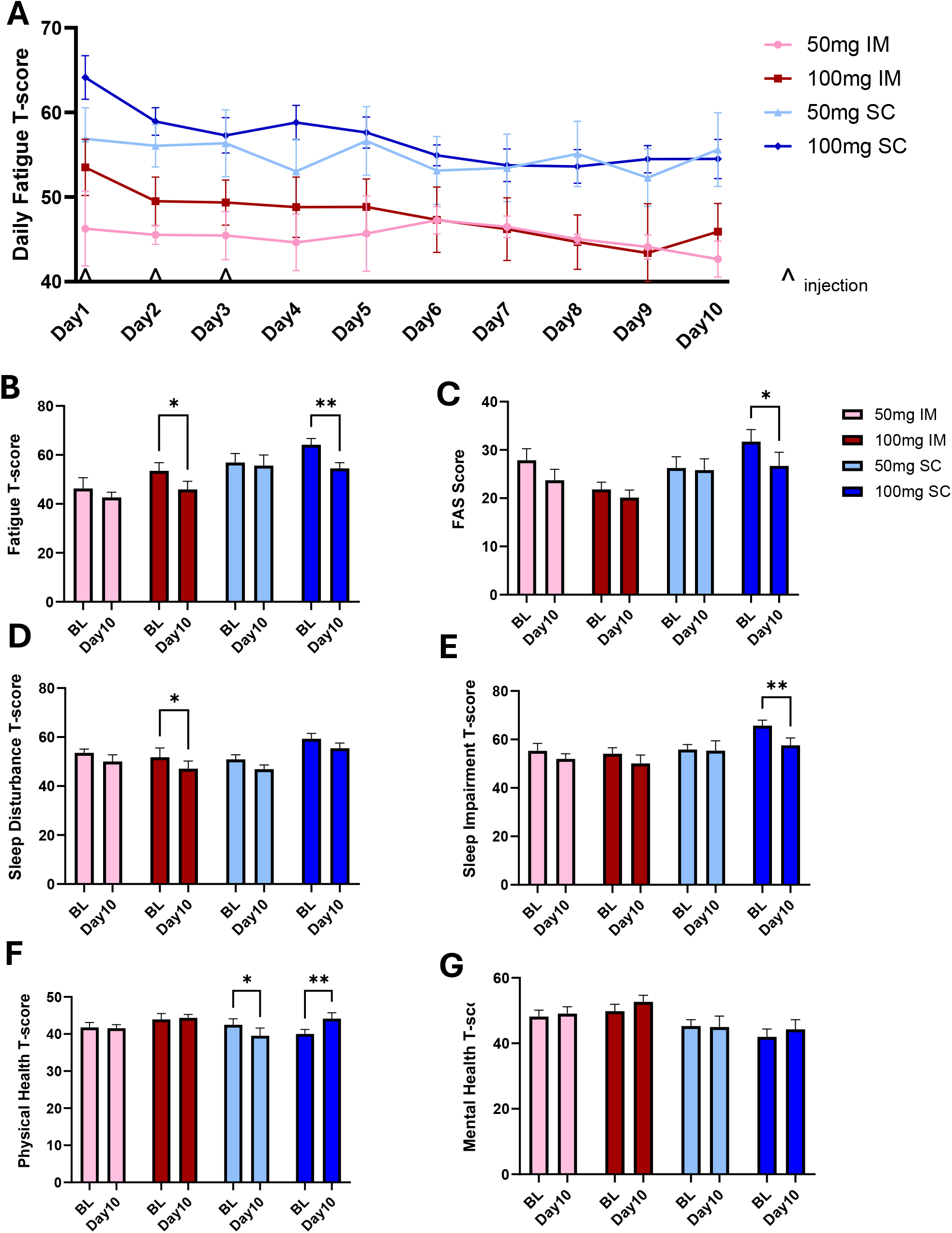
Participant Reported Outcomes for Fatigue, Sleep, and Global Physical and Mental Health. Perceived fatigue levels were assessed **A s B**) daily using the PROMIS Fatigue, Short Form 7b, and with the **C**) Fatigue Assessment Scale (FAS) at baseline (Day 1, Visit 1) and at the end of the study (Day 10, Visit 4). A decrease in either score indicates an improvement. **D s E**) Sleep quality was measured with the PROMIS® Sleep-Related Impairment and Sleep Disturbance questionnaire at baseline and on Day 10). A decrease in either score indicates an improvement. Quality of Life was assessed using the PROMIS® Global Health Scale Questionnaire at baseline and Day 10 and was categorized as **F)** global physical health and **G)** global mental health, where an increase in either score indicates improvement. The results from the PROMIS questionnaires are provided as T-scores. Values are reported as the group Average ±SEM. Statistical analysis included a mixed effects two-way ANOVA, with Tukey’s correction for multiple comparisons. * = p<0.05 and ** = p<0.01.

## Discussion

The safety and efficacy of parenteral NR administration remain relatively under-characterized. Therefore, the aim of this study was to compare the two doses and two routes of administration of NR, to support future protocols and product recommendations.

Injected NR at 50 and 100 mg doses, administered IM or SC for three consecutive days was well-tolerated, and did not result in treatment-related severe adverse events. Statistically significant changes in vital measurements and blood biomarkers, were not deemed clinically concerning by two separate healthcare providers, as they displayed no evidence of dose-dependent effects or a lack of recovery at the end of the washout period. However, it is recommended that such safety assessments continue to be used in larger, placebo-controlled studies, which may identify safety signals not observed in this study, due to its open-label and smaller study population size.

Oral administration of NR has resulted in modest reductions in SBP, that have not been validated in larger population studies^51^. An acute SBP reduction of approximately 10 mmHg was documented in the 50 mg SC group at 90 minutes post-injection. Though the extent of the reduction may appear clinically relevant, the changes were not sustained longitudinally. The effects were also inconsistent across groups, further complicating the interpretation of the isolated changes. Therefore, the physiological or clinical significance of these findings are unclear. Of note, longer-term oral supplementation with NAD+ precursor compounds have previously been demonstrated to exert anti-hypertensive effects, in some, but not all clinical studies ^51,85,86^. Further studies, with adequate statistical power, are required to determine the durability of parenteral NR’s effects on vascular health, and whether these effects vary based on baseline health characteristics.

Glucose, homocysteine, and hs-CRP were evaluated to identify potential acute safety signals in response to the injections. In both 50 mg groups, acute reductions in fasting blood glucose concentrations were observed following the first NR injection. While the magnitude of change was quantitatively modest and values remained within normal reference ranges, the findings may possibly signal an acute insulin-sensitizing or glucose flux-modulating effect of NR. These results are somewhat corroborated by Reyna et al., who identified very small reductions in glycosylated hemoglobin (HbA1C) values among a cohort of NR-IV users after a 30-day follow-up period^39^. When evaluating individual participant glucose responses, one participant in the 100 mg SC group presented markedly elevated glucose levels at baseline, i.e., >400 mg/dL likely secondary to unrecognized, uncontrolled hyperglycemia, throughout the study period. While this participant also experienced a drop in glucose following the first injection, their glucose levels continued to rise, even through baseline. However, no formal sensitivity analyses were conducted to quantify the influence of this observation, and thus its impact on group-level outcomes remain uncertain. The participant was retained in all primary analyses; no post-hoc exclusion or adjustments were made. Though an active diabetes diagnosis was grounds for exclusion in both trials, it may be prudent for future trials to employ both fasting plasma glucose and glycosylated hemoglobin (HbA1c) values to exclude participants with unrecognized uncontrolled glycemic dysregulation. While this single participant’s response was the most striking, when evaluating individual glucose patterns in the participants, there were other individuals that also had a similar pattern of drop in glucose with the first dose and an increase in glucose after subsequent injections and through the washout period. While their levels generally remained in the normal reference range, this pattern in some participants suggests heterogeneity in glucose response, and warrants monitoring, particularly in participants at risk of diabetes.

Participant provided adverse experience frequency did not appear dose-responsive for IM injections of NR, whereas for SC injections flushing/heat sensation, headache, and pain were more frequently reported with the higher dose. As this study is pilot in design, only one person reporting the experience for the lower dose and two participants for the higher dose is not sufficient to draw conclusions of a dose response. Important to note are the methodologic limitations that strongly limit the interpretability of these subjective experience observations – symptom data were collected via open-ended, unprompted self-report that were entirely qualitative in nature, rather than by a standardized, validated instrument. Thus, this introduces heterogeneity in how participants described their experiences and raises the likelihood that the apparent differences in adverse event frequencies may represent reporting style differences, rather than true pharmacologic differences. Further accentuating these limitations is the absence of formal severity grading. As such, it is therefore not possible to determine whether a report of “pain/discomfort” represented a brief, transient sensation or a more prolonged experience, nor to what extent the participant was affected. Therefore, frequency counts without severity context limits the clinical utility and interpretability of these data. Therefore, a more standardized approach evaluating participant experience for IM and SC injections warrants investigation in larger study populations before conclusions can be made.

This trial demonstrated NR injection tolerability and did not suggest safety signals. However, there are limitations to the study, including the exploratory, open-label study design and small sample size for each treatment arm. The absence of statistically significant adverse safety findings should be interpreted as a preliminary safety signal, not a definitive characterization of the long-term safety profile of injectable NR. The modest sample size precludes the ability to detect rare adverse events, or conclusively demonstrate differences in safety by relevant subgroups, e.g., age, sex, bodyweight.

Unfortunately, this trial did not incorporate a complete blood count, which would provide further evidence of safety. Variability in blood markers of glucose, homocysteine, and hsCRP suggest individual and unique responses to the injections, compromising general safety conclusions. Larger, powered studies, utilizing similar study designs, but employing full comprehensive metabolic panels and complete blood counts will help to develop population safety data, and reveal treatment-related changes in safety markers, if they exist. Additionally, preclinical, repeat-dose toxicology studies are needed to reveal similarities and variations between routes of NR administration and target organs of toxicity, which are currently unknown.

A formal standardized adverse event grading instrument such as the Common Terminology Criteria for Adverse Events (CTCAE) was not employed, which limits the ability to make severity-graded comparisons across groups, particularly with regards to the participant experience. Although these data collectively show that the injections are tolerable and do not result in safety signals, future research assessing participant experience using systematic and validated scales will better capture and quantify the participant experience.

Orally administered NR typically requires 10-14 days of continuous daily dosing to augment steady-state NAD+ levels in whole blood ^49,73,87^. The use and metabolism of oral NR by the gut bacteria likely contributes to this delay. Here, three days of injections did not result in statistically significant increases in whole blood NAD+ with each administration, likely due to heterogeneity in response and metabolism, which were demonstrated by the inter and intra-variability in time to maximum concentration with each injection. The time to maximum concentration (t_max_, after the baseline) ranged from 15 minutes to 24 hours, and such patterns varied by person with each injection. Thus, when NAD+ measurements were only taken at 90 minutes and 24 hours after the 2^nd^ and 3^rd^ injections, it is likely that the maximum concentration (C_max_) and t_max_ were not successfully captured for all participants for all three injections. This unanticipated variance precludes a conclusion regarding how repeated injectable NR augments whole blood NAD+, as such changes may not have been captured. However, an increasing number of participants did not have an observed C_max_ for the 2^nd^ and 3^rd^ injections, and thus the C_max_ for those participants was deemed 0. The daily average C_max_ was variable and is captured in Supplemental Figure 4. Significant heterogeneity in the pharmacokinetics of NAD+ was observed. Further research capturing multiple earlier and later time points to identify the daily t_max_ and C_max_ for individuals will help elucidate general population NAD+ pharmacokinetics. Additionally, individual NAD+ pharmacokinetic assessments for multiple days may be used to personalize treatment options and optimize dosing for injectable NR.

Globally, general fatigue prevalence amongst adults is 20.4% and the rate of fatigue that is experienced without an underlying medical diagnosis is ∼2.7 fold higher than fatigue associated with a medical condition^88^. NAD+ is an essential coenzyme required for energy metabolism and thus supports cellular energy^14,89^. Augmenting NAD+ at the cellular level, in theory may also reduce fatigue and support organism energy. To explore this theory, participants were required to have self-reported above average fatigue (FAS>22) for enrollment in the study. This inclusion criterion was implemented to develop general hypotheses around participant-reported outcomes. Over the ten days, while daily fatigue appeared to decrease slightly, the change was not statistically significant. However, when only assessing the Day 1 baseline and Day 10 changes, both the 100 mg IM and 100 mg SC groups showed statistically significant decreases in fatigue. This was strengthened by a statistically significant decrease in the FAS score for 100 mg SC, consistent with a possible reduction in fatigue that requires confirmation in a placebo-controlled trial.

Studies have shown that NAD+ precursor supplementation improves sleep quality in rodent models^90,91^, and improved sleep outcomes in a clinical trial conducted in Japan^92^. Sleep is a neurobiological state that is regulated by homeostatic and circadian rhythms^82^, both of which are impacted by NAD+^93,94^. The domain of sleep disturbance encompasses perceptions of sleep quality, time to fall asleep and restlessness, where values of 55 or greater indicate a potential impairment in this area. While all treatment groups demonstrated a moderate reduction in sleep disturbance, the 100 mg IM group resulted in a statistically significant decrease. The sleep impairment domain of the PROMIS questionnaire assesses how sleep problems impact wakefulness including alertness, daytime sleepiness, concentration, irritability, and accomplishing tasks^95^. Similar to sleep disturbance, a score above 55 indicates sleep impairment that is greater than the general U.S. population, and scores of 60.0 to 69.9 indicate moderate sleep impairment. Only the 100 mg SC group had an average sleep impairment score above 60, and this group was the only group to show a statistically significant decrease with treatment. While it is interesting that the different routes of administration resulted in different sleep domain benefits, such results will need to be validated in larger, powered, and placebo-controlled studies to support sleep claims.

Quality of Life was measured with two domains, global physical health and global mental health. The average score for these two domains in the United States is 50, and scores below 50 indicate various degrees of impairment, with lower scores indicating increased severity. The global physical health score assesses perceived general physical health, ability to complete everyday activities, average pain intensity and fatigue^84^. Interestingly, the 50 mg SC dose statistically significantly decreased the physical health score, while 100 mg SC resulted in a statistically significant improvement. The global mental health domain evaluates the perception of overall quality of life, emotional and psychological well-being, and social engagement satisfaction. Including relationships. A score of 50 represents the US average, and higher scores indicate a better perception of mental and emotional well-being. Over the 10 days, the global mental health score did not statistically change for any of the treatment groups though the 100 mg IM and 100 mg SC groups had slight increases, that should be evaluated in longer and larger trials to detect an actual signal.

The absence of a placebo arm limits causal inferences regarding injectable NR; as some of the observed changes may have arisen due to confounders, such as background lifestyle factors, random variation, temporal effects, or regression to the mean. Without a placebo control arm, within-group changes cannot be attributed to the intervention as they cannot be distinguished from factors unrelated to the intervention, such as time-of-day, fasting state, regression to the mean, or repeated venipuncture artifact. The open-label nature of the study also introduces the potential for expectancy bias that may influence PRO. All reported within-arm changes from baseline should be considered descriptive only, and support hypothesis generation. Values that were determined to be statistically significant are statistical only, not causal, and require validation in a larger, powered, placebo-controlled study.

## Conclusion

NR was generally well-tolerated and was not associated with safety risks in this study population. These preliminary data suggest potential acute benefits of NR injections on transient increases in whole blood NAD+ in most participants, fatigue, sleep, and global physical health. Future clinical trials are needed to validate findings and support claims/indications for injectable NR.

## Data Availability

All data produced in the present study are available upon reasonable request to the corresponding author.

## Acknowledgements

- The team is extremely grateful to the study participants for contributing to this important scientific advancement.
- Thank you to Anne Russ for going above and beyond, for your attention to detail, and <u>for</u> learning on the fly to get this trial completed and <u>the</u> manuscript to the finish line.
- Thank you to Halland Chen, MD, for serving as the PI, supporting study conceptualization, the IRB process, and supervising the clinical trial.
- Thank you to the staff at IHM for your contributions towards managing and conducting the clinical trial.
- Thank you Kris Fishman, Eric Huynh, and the team at Wells Pharmacy for compounding Niagen®Plus and helping identify a pathway to supply the material.
- Thank you, Chris Meletis, ND for reviewing the blood biomarker and vitals data.
- Thank you Albert Rembert for supervising clinical site visits and providing quality assurance and control guidelines.
- Thank you, Anthony Low, Esq., for handling all the legal agreements to make the studies happen. You are such a valuable team member.
- Thank you Niagen Bioscience TruCrew and ELT for supporting our external research program and Raising the BAR.
- Thank you to Dr. Marc Abreu, MD, for serving as the independent medical monitor for this trial and for meeting with YNE and JK to discuss the importance and relevance of this clinical trial.

## Statement of Contribution

- All authors were required to review and provide edits on the manuscript. Additionally, all authors were required to approve the final version prior to submission.
- YNE facilitated conceptualization, study initiation, and sponsor-side project management during the in-clinic and early data analysis portion of the study. She also contributed to the development of study protocols, data analysis and interpretation, data visualization, manuscript writing, editing and final manuscript review.
- JK contributed sponsor-side project management for the completion of this manuscript. He also was involved in data interpretation, manuscript writing, editing, and final manuscript review.
- AR served as the clinical trial manager, was responsible for final protocol development, clinical trial management, data collection, data analysis and interpretation, and final review of the manuscript.
- SS served as the lead statistician and contributed to data analysis and interpretation, manuscript editing and final manuscript review.
- FH contributed to data analysis, manuscript writing, editing, and final manuscript review.
- TA contributed to data collection, manuscript editing, and final manuscript review.
- HC served as the PI for the study, contributing to conceptualization, study supervision, securing resources, and final manuscript editing and approval.
- AS contributed to conceptualization, securing resources, and final manuscript editing and approval.

## Declaration of Generative AI Use

Generative AI was not used in the development of this manuscript.

