## Supplementary for "Part 1 of a 2-part, open-label clinical trial evaluating repeated nicotinamide riboside parenteral administration: 3 consecutive days of intramuscular and subcutaneous injections"

### Part 1 Manuscript Supplementary Tables and Figures

Supplemental Table 1. Participant Vitals

| Vital Parameter | Treatment Group | Group Average ±SEM | Day 1, 1st Inj. V1 BL | Day 1 V1 90 min | Δ V1 BL to 90 min post injection Statistical Significance & change | Day 1 V1 180 min | ΔV1 BL to V1 180 min Statistical Significance & change | Day 2 V2 90 min | Day 3 V3 90 min | Day 10 V4 BL | ΔV1 BL to V4 BL Statistical Significance & change |
|---|---|---|---|---|---|---|---|---|---|---|---|
| SBP, mmHG | 50 mg IM | Average | 127.71 | 120.57 | ns | 121.71 | ns | 118.57 | 115.33 | 116.43 | ns |
| | | ± SEM | 5.91 | 5.88 | | 5.86 | | 5.37 | 4.82 | 5.32 | |
| | 50 mg SC | Average | 130.70 | 120.10 | p=0.011 ↓ (8.11%) | 123.80 | ns | 117.60 | 125.30 | 123.80 | ns |
| | | ± SEM | 6.14 | 4.74 | | 4.81 | | 4.92 | 6.35 | 4.95 | |
| | 100 mg IM | Average | 129.55 | 131.27 | ns | 128.18 | ns | 130.18 | 122.73 | 127.73 | ns |
| | | ± SEM | 4.97 | 4.60 | | 4.88 | | 4.12 | 3.87 | 5.43 | |
| | 100 mg SC | Average | 129.33 | 128.33 | ns | 127.56 | ns | 124.56 | 123.67 | 119.67 | ns |
| | | ± SEM | 2.84 | 4.42 | | 3.54 | | 3.87 | 3.45 | 4.90 | |
| DBP, mmHG | 50 mg IM | Average | 80.00 | 77.71 | ns | 75.14 | ns | 79.14 | 76.00 | 75.43 | ns |
| | | ± SEM | 3.73 | 4.63 | | 2.31 | | 3.60 | 3.40 | 3.34 | |
| | 50 mg SC | Average | 76.00 | 70.90 | ns | 70.89 | ns | 69.40 | 75.20 | 75.70 | ns |
| | | ± SEM | 3.09 | 2.98 | | 2.50 | | 3.09 | 4.21 | 3.13 | |
| | 100 mg IM | Average | 81.55 | 82.82 | ns | 77.45 | ns | 81.36 | 82.73 | 81.55 | ns |
| | | ± SEM | 2.84 | 3.05 | | 3.65 | | 4.31 | 2.24 | 5.46 | |
| | 100 mg SC | Average | 78.56 | 78.11 | ns | 74.11 | ns | 78.89 | 77.78 | 79.56 | ns |
| | | ± SEM | 5.94 | 4.06 | | 4.80 | | 5.88 | 4.68 | 4.11 | |
| HR, bpm | 50 mg IM | Average | 71.43 | 71.00 | ns | 69.43 | ns | 72.43 | 70.83 | 73.43 | ns |
| | | ± SEM | 6.00 | 5.48 | | 5.62 | | 1.90 | 2.39 | 4.48 | |
| | 50 mg SC | Average | 64.70 | 65.00 | ns | 72.30 | ns | 61.80 | 65.10 | 67.40 | ns |
| | | ± SEM | 2.94 | 2.57 | | 2.77 | | 2.64 | 2.85 | 3.11 | |
| | 100 mg IM | Average | 70.00 | 67.82 | ns | 80.27 | p=0.006 ↑(14.67%) | 69.27 | 67.18 | 70.91 | ns |
| | | ± SEM | 1.93 | 1.41 | | 4.00 | | 3.48 | 2.34 | 2.44 | |
| | 100 mg SC | Average | 71.44 | 69.22 | ns | 77.00 | ns | 77.22 | 70.44 | 78.78 | ns |
| | | ± SEM | 3.03 | 2.56 | | 4.42 | | 4.46 | 1.89 | 3.38 | |
| RR, brpm | 50 mg IM | Average | 17.29 | 16.17 | ns | 16.29 | p=0.027 ↓(5.78%) | 16.00 | 16.00 | 16.29 | ns |
| | | ± SEM | 0.36 | 0.17 | | 0.29 | | 0.00 | 0.00 | 0.29 | |
| | 50 mg SC | Average | 15.10 | 15.60 | ns | 15.60 | ns | 15.10 | 16.40 | 16.00 | ns |

| | | | | | | | | | | | |
|---|---|---|---|---|---|---|---|---|---|---|---|
| | | ± SEM | 0.64 | 0.27 | | 0.27 | | 0.48 | 0.27 | 0.00 | |
| | 100 mg IM | Average | 15.91 | 15.36 | ns | 15.82 | ns | 15.36 | 16.00 | 15.64 | ns |
| | | ± SEM | 0.34 | 0.43 | | 0.33 | | 0.34 | 0.00 | 0.24 | |
| | 100 mg SC | Average | 15.50 | 14.67 | ns | 15.33 | ns | 15.00 | 15.33 | 15.56 | ns |
| | | ± SEM | 0.50 | 0.73 | | 0.75 | | 0.53 | 0.47 | 0.29 | |
| SpO2 (%) | 50 mg IM | Average | 98.86 | 98.43 | ns | 98.71 | ns | 98.00 | 98.33 | 98.29 | ns |
| | | ± SEM | 0.59 | 0.57 | | 0.61 | | 0.53 | 0.61 | 0.52 | |
| | 50 mg SC | Average | 98.70 | 98.90 | ns | 97.90 | ns | 98.90 | 99.00 | 98.60 | ns |
| | | ± SEM | 0.42 | 0.28 | | 0.43 | | 0.31 | 0.30 | 0.43 | |
| | 100 mg IM | Average | 98.55 | 98.18 | ns | 98.18 | ns | 98.82 | 98.55 | 98.27 | ns |
| | | ± SEM | 0.49 | 0.33 | | 0.44 | | 0.40 | 0.47 | 0.43 | |
| | 100 mg SC | Average | 99.67 | 98.00 | 0.013 ↓(1.68%) | 98.67 | p=0.022 ↓(1.00%) | 98.89 | 98.33 | 98.33 | ns |
| | | ± SEM | 0.17 | 0.47 | | 0.47 | | 0.35 | 0.65 | 0.55 | |
| Temp, °F | 50 mg IM | Average | 97.89 | 98.20 | ns | 98.67 | ns | 97.86 | 98.12 | 98.00 | ns |
| | | ± SEM | 0.24 | 0.26 | | 0.19 | | 0.10 | 0.48 | 0.22 | |
| | 50 mg SC | Average | 98.21 | 98.05 | ns | 97.77 | ns | 98.09 | 97.41 | 97.79 | ns |
| | | ± SEM | 0.15 | 0.46 | | 0.47 | | 0.25 | 0.23 | 0.15 | |
| | 100 mg IM | Average | 97.57 | 98.07 | ns | 98.48 | ns | 97.99 | 97.13 | 97.89 | ns |
| | | ± SEM | 0.24 | 0.27 | | 0.35 | | 0.22 | 0.28 | 0.31 | |
| | 100 mg SC | Average | 98.21 | 98.54 | ns | 98.87 | ns | 99.04 | 97.94 | 98.11 | ns |
| | | ± SEM | 0.41 | 0.22 | | 0.22 | | 0.33 | 0.34 | 0.21 | |

Supplemental Table 2. Blood biomarkers for safety assessment

| Blood Biomarker | Treatment Group | V1 Day 1 BL Avg ±SEM | V1, 90 min post injection Avg ±SEM | Δ V1 BL to 90 min post injection Statistical Significance & change | V2 Day 2 Post Inj. 2 | V3 Day 3 Post Inj. 3 | V4 Day 10 Post Washout | Δ V1 BL to V4 BL Statistical Significance & change |
|---|---|---|---|---|---|---|---|---|
| **Albumin** | 50 mg IM | 4.42±0.11 | 4.45±0.09 | ns | 4.53±0.10 | 4.46±0.09 | 4.55±0.10 | p=0.039 ↑ (+2.7%) |
| | 50 mg SC | 4.57±0.14 | 4.46±0.09 | ns | 4.52±0.11 | 4.44±0.12 | 4.31±0.11 | p=0.040 ↓ (-5.7%) |
| | 100 mg IM | 4.54±0.08 | 4.51±0.09 | ns | 4.51±0.08 | 4.53±0.06 | 4.34±0.09 | p=0.006 ↓ (-4.5%) |
| | 100 mg SC | 4.40±0.08 | 4.47±0.10 | ns | 4.44±0.13 | 4.53±0.14 | 4.50±0.14 | ns |
| **Alkaline Phosphatase** | 50 mg IM | 72.59±7.78 | 72.81±7.38 | ns | 73.94±7.99 | 71.97±7.98 | 74.47±8.36 | ns |
| | 50 mg SC | 57.96±7.05 | 57.87±7.50 | ns | 56.97±7.18 | 55.67±6.94 | 57.46±6.80 | ns |
| | 100 mg IM | 63.41±5.88 | 62.06±5.92 | ns | 63.91±6.27 | 63.81±6.63 | 61.95±6.81 | ns |
| | 100 mg SC | 66.57±3.79 | 66.48±3.74 | ns | 66.49±2.59 | 66.80±3.00 | 70.33±3.61 | ns |
| **ALT** | 50 mg IM | 29.63±9.25 | 28.84±8.85 | ns | 28.19±9.14 | 27.10±9.49 | 23.49±5.98 | ns |
| | 50 mg SC | 27.77±7.23 | 27.09±7.22 | ns | 25.85±6.52 | 23.41±4.64 | 25.34±4.08 | ns |
| | 100 mg IM | 34.99±5.33 | 34.58±5.36 | ns | 37.15±6.96 | 36.16±6.59 | 32.73±5.17 | ns |
| | 100 mg SC | **42.84±18.10** | **42.60±18.13** | ns | **42.06±17.93** | **41.82±17.39** | **50.76±26.10** | ns |
| **AST** | 50 mg IM | 24.77±3.59 | 23.94±3.28 | ns | 23.49±3.86 | 22.93±3.86 | 22.37±2.72 | ns |
| | 50 mg SC | 26.27±4.29 | 25.70±4.60 | ns | 23.11±3.60 | 21.87±1.83 | 29.21±9.11 | ns |
| | 100 mg IM | 30.36±3.73 | 30.28±3.81 | ns | 31.41±4.25 | 29.81±4.03 | 26.65±2.67 | ns |
| | 100 mg SC | 31.76±9.73 | 31.58±9.62 | ns | 30.59±9.27 | 29.92±8.39 | 35.84±14.74 | ns |
| **Bili Total** | 50 mg IM | 0.47±0.05 | 0.52±0.04 | ns | 0.47±0.06 | 0.51±0.03 | 0.46±0.05 | ns |
| | 50 mg SC | 0.54±0.05 | 0.55±0.04 | ns | 0.45±0.04 | 0.53±0.04 | 0.43±0.04 | ns |
| | 100 mg IM | 0.54±0.09 | 0.58±0.08 | ns | 0.59±0.10 | 0.60±0.10 | 0.53±0.09 | ns |
| | 100 mg SC | 0.61±0.07 | 0.63±0.07 | ns | 0.55±0.07 | 0.55±0.06 | 0.59±0.05 | ns |

| | | | | | | | | |
|---|---|---|---|---|---|---|---|---|
| **BUN** | 50 mg IM | 12.87±1.26 | 12.06±1.29 | p<0.001 ↓ (-6.3%) | 14.69±2.04 | 12.77±2.18 | 13.70±1.42 | ns |
| | 50 mg SC | 12.88±1.49 | 12.21±1.48 | p=0.019 ↓ (-5.2%) | 13.64±0.98 | 13.16±1.18 | 13.10±1.37 | ns |
| | 100 mg IM | 16.18±1.99 | 14.38±1.54 | ns | 14.98±2.02 | 14.39±1.82 | 15.23±1.52 | ns |
| | 100 mg SC | 13.22±2.23 | 12.77±2.15 | ns | 12.83±1.60 | 12.34±1.48 | 13.49±1.49 | ns |
| **Calcium** | 50 mg IM | 9.37±0.12 | 9.50±0.09 | ns | 9.71±0.12 | 9.51±0.10 | 9.45±0.09 | ns |
| | 50 mg SC | 9.56±0.10 | 9.46±0.09 | ns | 9.48±0.12 | 9.45±0.13 | 9.41±0.13 | ns |
| | 100 mg IM | 9.52±0.09 | 9.52±0.13 | ns | 9.58±0.11 | 9.58±0.10 | 9.42±0.09 | ns |
| | 100 mg SC | 9.43±0.09 | 9.47±0.08 | ns | 9.54±0.14 | 9.57±0.11 | 9.59±0.15 | ns |
| **Carbon Dioxide** | 50 mg IM | 21.59±0.45 | 21.87±0.62 | ns | 21.99±0.62 | 22.56±0.44 | 22.21±0.36 | ns |
| | 50 mg SC | 22.19±0.59 | 21.62±0.73 | ns | 22.96±0.81 | 22.20±0.76 | 23.17±0.50 | p=0.048 ↑ (+4.4%) |
| | 100 mg IM | 22.47±0.75 | 23.15±0.49 | ns | 23.13±0.63 | 22.95±0.53 | 22.58±0.61 | ns |
| | 100 mg SC | 20.94±0.26 | 21.54±0.65 | ns | 22.70±0.46 | 22.61±0.33 | 21.69±0.61 | ns |
| **Chloride** | 50 mg IM | 103.54±0.62 | 103.01±0.68 | ns | 102.23±0.40 | 102.71±0.86 | 103.84±0.55 | ns |
| | 50 mg SC | 103.07±0.66 | 103.48±0.38 | ns | 103.66±0.49 | 103.70±0.51 | 104.26±0.35 | ns |
| | 100 mg IM | 103.01±0.60 | 102.68±0.61 | ns | 103.56±0.67 | 103.41±0.50 | 103.95±0.65 | p=0.013 ↑ (+0.9%) |
| | 100 mg SC | 102.32±0.88 | 102.38±0.77 | ns | 102.61±1.08 | 103.09±0.99 | 103.00±0.94 | ns |
| **Creatinine** | 50 mg IM | 0.77±0.08 | 0.76±0.08 | ns | 0.78±0.08 | 0.75±0.08 | 0.76±0.08 | ns |
| | 50 mg SC | 0.80±0.07 | 0.78±0.06 | ns | 0.78±0.06 | 0.78±0.06 | 0.76±0.05 | ns |
| | 100 mg IM | **0.97±0.08** | **0.95±0.08** | ns | **0.94±0.08** | **0.94±0.07** | **0.95±0.07** | ns |
| | 100 mg SC | **0.94±0.08** | **0.95±0.07** | ns | **0.93±0.07** | **0.95±0.07** | **0.99±0.08** | p=0.003 ↑ (+5.4%) |
| **Glucose** | 50 mg IM | 90.47±1.39 | 83.96±1.90 | p=0.003 ↓ (-7.2%) | 87.56±2.04 | 85.21±2.00 | 89.91±3.19 | ns |
| | 50 mg SC | 90.41±2.47 | 85.71±1.48 | p=0.009 ↓ (-5.2%) | 87.96±2.10 | 86.90±2.14 | 92.53±2.80 | ns |

| | | | | | | | | |
|---|---|---|---|---|---|---|---|---|
| | 100 mg IM | 91.25±2.01 | 88.25±1.93 | p=0.041 ↓ (-3.3%) | 88.05±1.54 | 86.94±2.41 | 89.35±2.13 | ns |
| | 100 mg SC | **115.76±28.68** | **108.86±25.21** | ns | **117.28±28.25** | **120.91±34.18** | **129.62±37.61** | ns |
| **Hematocrit** | 50 mg IM | 40.83±1.50 | 41.29±0.67 | ns | 42.07±1.26 | 41.26±0.99 | 41.59±1.09 | ns |
| | 50 mg SC | 40.99±0.79 | 40.73±0.82 | ns | 40.16±0.70 | 40.27±0.32 | **39.74±0.74** | p=0.047 ↓ (-3.0%) |
| | 100 mg IM | 43.95±1.52 | 43.45±1.50 | ns | 43.20±1.49 | 43.93±1.67 | 42.46±1.58 | p=0.026 ↓ (-3.4%) |
| | 100 mg SC | 41.77±1.10 | 41.71±1.15 | ns | 41.57±1.15 | 41.39±0.89 | 41.96±1.01 | ns |
| **Hemoglobin** | 50 mg IM | **13.41±0.46** | 13.73±0.28 | ns | 14.07±0.48 | 13.80±0.37 | 13.73±0.41 | ns |
| | 50 mg SC | **13.32±0.29** | **13.26±0.28** | ns | **13.21±0.29** | **13.19±0.22** | **12.96±0.28** | p=0.047 ↓ (-2.7%) |
| | 100 mg IM | 14.62±0.57 | 14.50±0.57 | ns | 14.46±0.56 | 14.50±0.57 | 14.08±0.57 | p=0.002 ↓ (-3.7%) |
| | 100 mg SC | 13.77±0.37 | 13.90±0.36 | ns | 13.88±0.38 | **13.69±0.37** | 13.91±0.34 | ns |
| **MCH** | 50 mg IM | 30.83±0.47 | 30.91±0.42 | ns | 30.76±0.45 | 30.47±0.52 | 30.61±0.63 | ns |
| | 50 mg SC | 29.56±0.54 | 29.63±0.45 | ns | 29.73±0.54 | 29.67±0.48 | 29.61±0.46 | ns |
| | 100 mg IM | 30.02±0.59 | 29.93±0.58 | ns | 29.93±0.58 | 29.85±0.62 | 29.85±0.58 | ns |
| | 100 mg SC | 30.66±0.66 | 30.89±0.69 | ns | 30.78±0.68 | 30.31±0.64 | 30.66±0.62 | ns |
| **MCHC** | 50 mg IM | 32.87±0.51 | 33.23±0.34 | ns | 33.43±0.46 | 33.46±0.31 | 33.00±0.32 | ns |
| | 50 mg SC | 32.48±0.25 | 32.55±0.24 | ns | 32.87±0.30 | 32.73±0.33 | 32.61±0.31 | ns |
| | 100 mg IM | 33.21±0.22 | 33.33±0.28 | ns | 33.45±0.27 | 32.99±0.25 | 33.12±0.28 | ns |
| | 100 mg SC | 32.97±0.38 | 33.36±0.40 | p=0.032 ↑ (+1.2%) | 33.41±0.33 | 33.06±0.33 | 33.16±0.33 | ns |
| **MCV** | 50 mg IM | **93.74±0.70** | **93.01±0.85** | ns | 91.96±0.94 | 91.16±1.30 | **92.67±1.49** | ns |
| | 50 mg SC | 90.89±1.27 | 90.98±1.16 | ns | 90.36±1.12 | 90.63±1.13 | 90.84±1.14 | ns |
| | 100 mg IM | 90.40±1.64 | 89.83±1.62 | ns | 89.48±1.57 | 90.43±1.84 | 90.05±1.55 | ns |
| | 100 mg SC | **92.91±1.51** | **92.57±1.43** | ns | 92.07±1.39 | 91.74±1.62 | **92.39±1.58** | ns |
| **Platelet Count** | 50 mg IM | 278.43±38.73 | 282.71±28.67 | ns | 296.14±31.97 | 295.14±29.61 | 321.43±48.16 | ns |
| | 50 mg SC | 298.00±25.86 | 285.50±24.13 | ns | 286.70±24.88 | 284.30±22.49 | 283.30±21.86 | ns |
| | 100 mg IM | 253.36±15.65 | 254.00±14.16 | ns | 257.64±16.87 | 259.27±17.65 | 268.82±18.53 | ns |

| | | | | | | | | |
|---|---|---|---|---|---|---|---|---|
| | 100 mg SC | 282.44±20.62 | 279.44±19.96 | ns | 290.89±21.42 | 288.56±24.33 | 286.44±24.59 | ns |
| **Potassium** | 50 mg IM | 4.35±0.11 | 4.29±0.14 | ns | 4.40±0.19 | 4.24±0.15 | 4.43±0.14 | ns |
| | 50 mg SC | 4.32±0.06 | 4.32±0.09 | ns | 4.45±0.14 | 4.33±0.12 | 4.33±0.08 | ns |
| | 100 mg IM | 4.39±0.10 | 4.37±0.09 | ns | 4.43±0.08 | 4.34±0.11 | 4.35±0.08 | ns |
| | 100 mg SC | 4.49±0.10 | 4.39±0.08 | ns | 4.42±0.10 | 4.50±0.08 | 4.56±0.10 | ns |
| **Protein Total** | 50 mg IM | 7.10±0.24 | 7.12±0.19 | ns | 7.33±0.25 | 7.15±0.24 | 7.12±0.27 | ns |
| | 50 mg SC | 7.23±0.16 | 7.05±0.11 | ns | 7.01±0.08 | 6.89±0.12 | 6.88±0.11 | ns |
| | 100 mg IM | 7.08±0.07 | 7.14±0.09 | ns | 7.20±0.12 | 7.16±0.08 | 6.87±0.13 | ns |
| | 100 mg SC | 6.78±0.08 | 6.88±0.12 | ns | 6.87±0.16 | 6.97±0.14 | 7.00±0.18 | ns |
| **RBC** | 50 mg IM | 4.36±0.18 | 4.44±0.10 | ns | 4.58±0.15 | 4.53±0.12 | 4.50±0.16 | ns |
| | 50 mg SC | 4.51±0.08 | 4.48±0.09 | ns | 4.45±0.09 | 4.45±0.07 | 4.38±0.09 | p=0.029 ↓ (-3.0%) |
| | 100 mg IM | 4.88±0.18 | 4.85±0.17 | ns | 4.84±0.17 | 4.87±0.19 | 4.73±0.20 | p=0.020 ↓ (-2.9%) |
| | 100 mg SC | 4.51±0.17 | 4.52±0.18 | ns | 4.53±0.17 | 4.53±0.16 | 4.56±0.17 | ns |
| **Sodium** | 50 mg IM | 139.03±0.58 | 138.77±0.63 | ns | 138.59±0.28 | 139.26±0.52 | 139.79±1.01 | ns |
| | 50 mg SC | 138.98±0.63 | 139.16±0.53 | ns | 139.14±0.66 | 139.13±0.44 | 140.39±0.42 | p=0.027 ↑ (+1.0%) |
| | 100 mg IM | 139.62±0.58 | 139.14±0.55 | ns | 140.36±0.62 | 139.84±0.54 | 140.45±0.77 | ns |
| | 100 mg SC | 137.63±0.57 | 138.26±0.67 | p=0.030 ↑ (+0.5%) | 138.89±0.77 | 139.36±0.84 | 138.72±0.83 | ns |
| **WBC** | 50 mg IM | 5.78±0.33 | 6.52±0.49 | p=0.041 ↑ (+12.8%) | 6.24±0.44 | 6.56±0.46 | 6.59±0.70 | ns |
| | 50 mg SC | 5.46±0.53 | 5.61±0.48 | ns | 5.60±0.42 | 5.59±0.39 | 5.53±0.43 | ns |
| | 100 mg IM | 5.94±0.55 | 5.99±0.50 | ns | 6.24±0.63 | 5.63±0.35 | 5.31±0.38 | ns |
| | 100 mg SC | 5.52±0.37 | 5.64±0.41 | ns | 5.65±0.38 | 5.91±0.52 | 5.94±0.60 | ns |

**BOLD** indicates values outside of normal reference ranges for that parameter

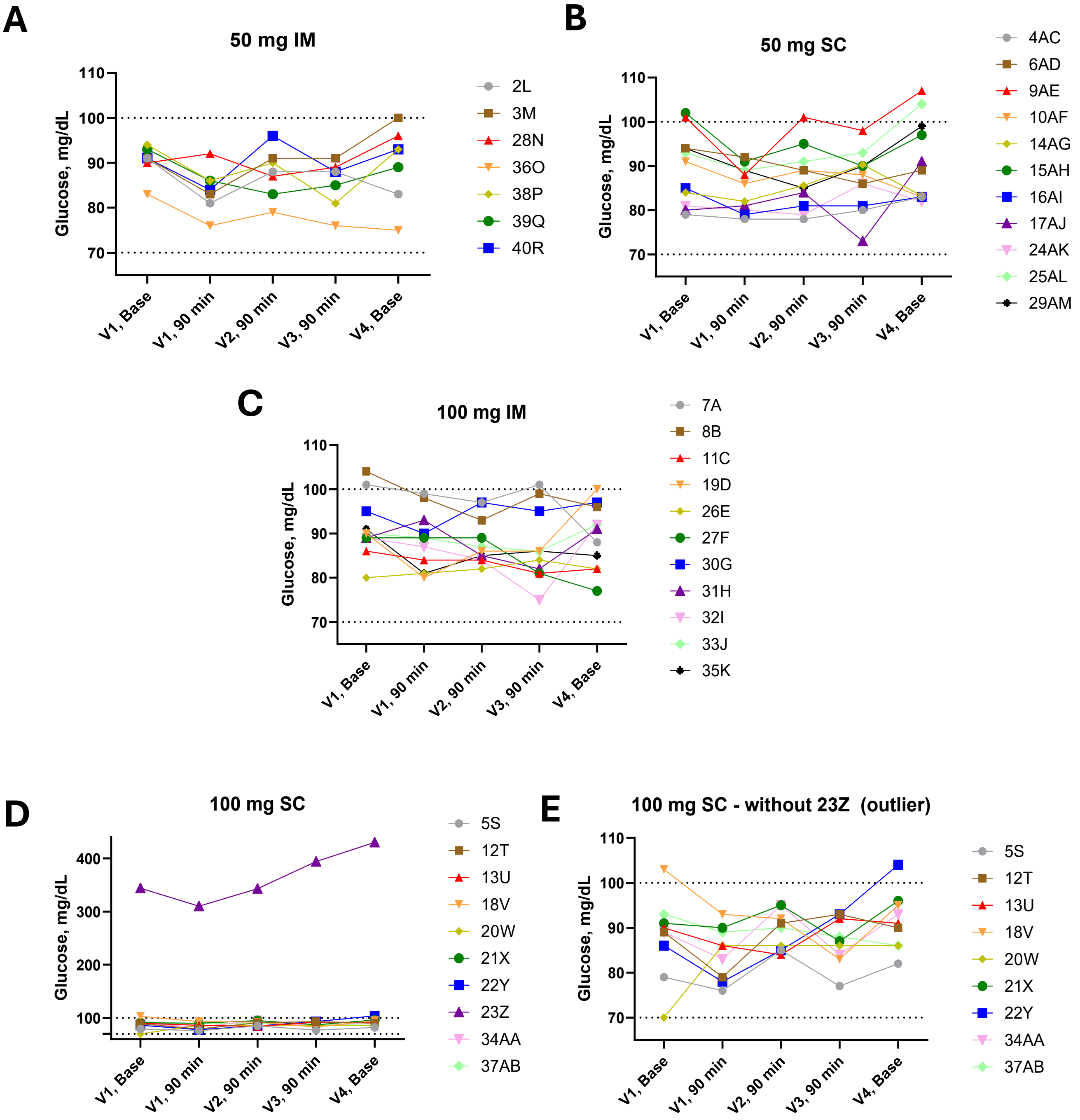


**Supplementary Figure 1. Individual longitudinal changes in serum glucose by dose and route of administration.** Participant's glucose results are categorized by treatment group for **A**) 50 mg IM, **B**) 50 mg SC, **C**) 100 mg IM, **D**) 100 mg SC and E) 100 mg SC without the outlier. The normal reference range of 70 to 100 mg/dL for fasted glucose is indicated by the dashed lines. The injections did not illicit general group direction responses in glucose changes with each administration.

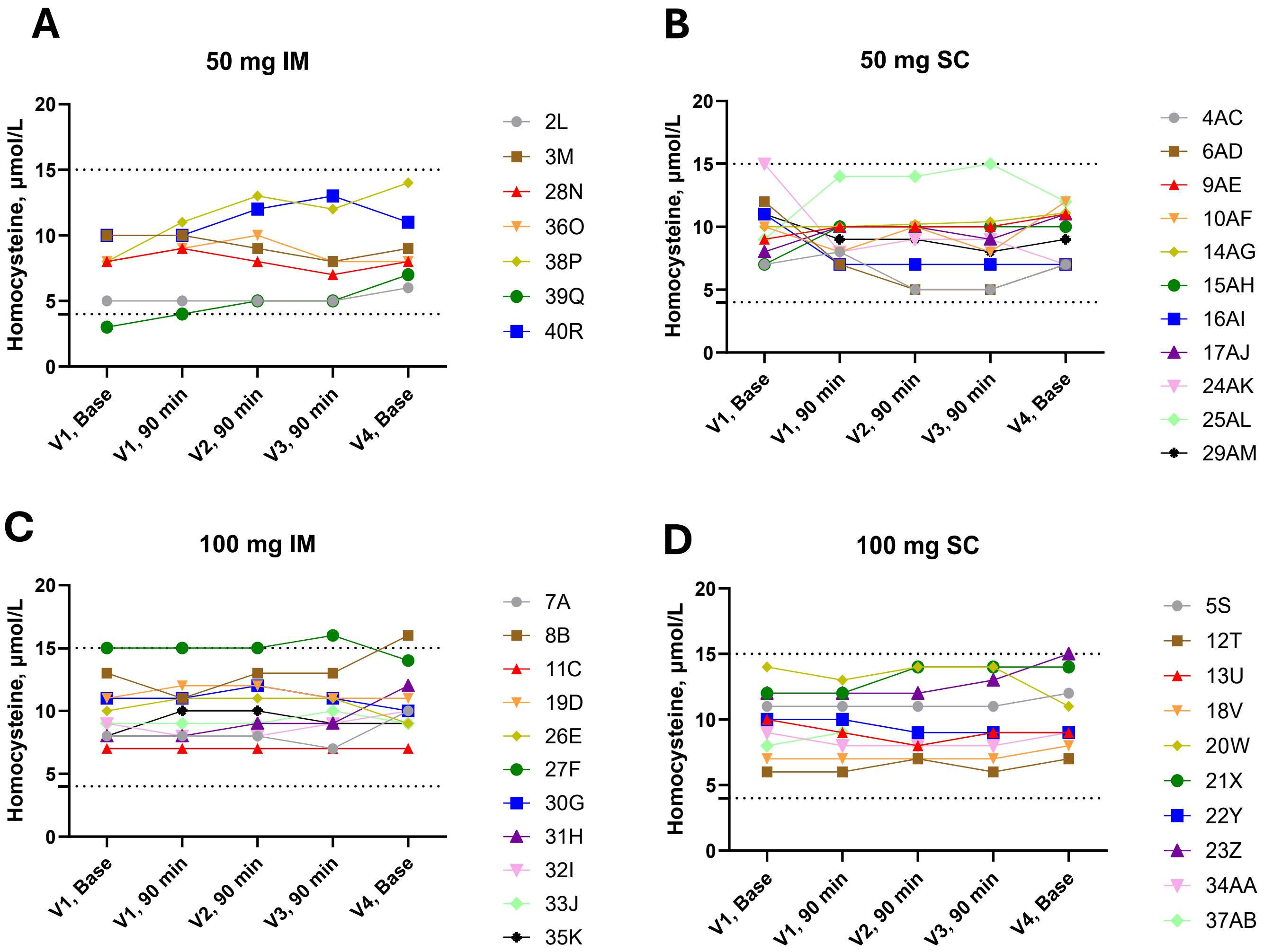


**Supplemental Figure 2. Individual longitudinal changes in homocysteine by dose and route of administration.** Participant's homocysteine results are categorized by treatment group for **A**) 50 mg IM, **B**) 50 mg SC, **C**) 100 mg IM, **D**) 100 mg SC. The normal reference range of 4 to 15 μmol/L is indicated by the dashed lines on each figure. The injections did not illicit general group directional changes in homocysteine in response to the injections.

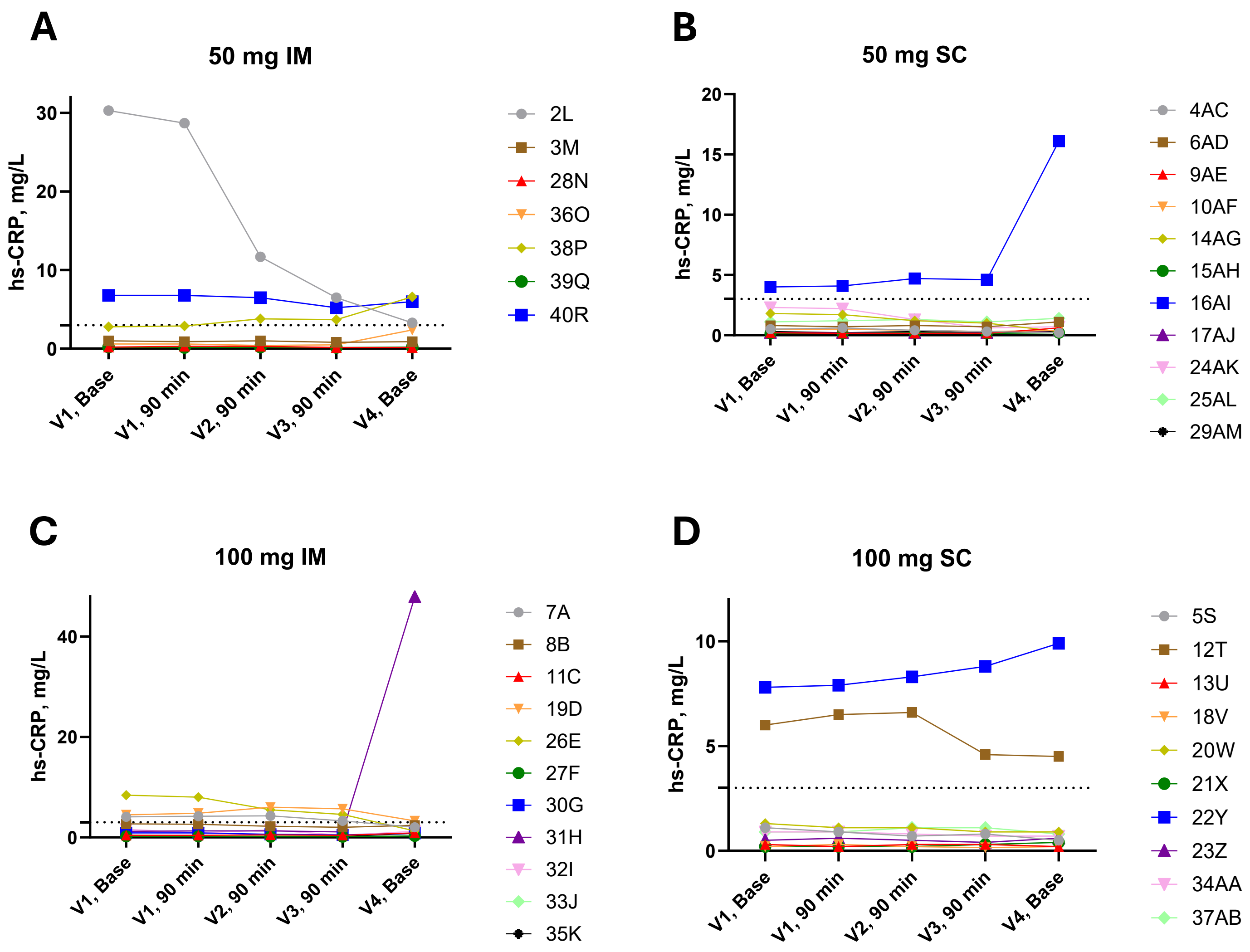


**Supplemental Figure 3. Individual longitudinal changes in hsCRP by dose and route of administration.** Participant's hsCRP results are categorized by treatment group for **A**) 50 mg IM, **B**) 50 mg SC, **C**) 100 mg IM, **D**) 100 mg SC as a clinical indicator of systemic inflammation. The normal reference range of <3.0 mg/L for hsCRP is indicated by the dashed lines on each figure. The injections did not illicit general group directional changes in hs-CRP in response to the injections.

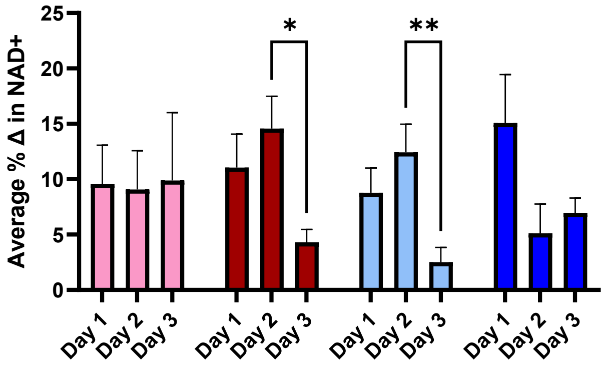


**Supplemental Figure 4. Average daily NAD+ increase**. The percent increase in NAD+ from each day's baseline to individual $C_{max}$ with each injection. If a $C_{max}$ was not observed, the percent increase was listed as 0%. Values are reported as the group average ±SEM. Statistical analysis included a mixed effects two-way ANOVA, with Tukey's correction for multiple comparisons. * = $p<0.05$ and ** = $p<0.01$.

Supplemental Table 3. Percentage of participants in each treatment group that experienced each adverse response with one or more of the injections.

| **Participant reported injection experience** | **50 mg IM** | **50 mg SC** | **100 mg IM** | **100 mg SC** | **50 mg** | **100 mg** | **IM** | **SC** |
|---|---|---|---|---|---|---|---|---|
| Bruising | 0.0 | 0.0 | 0.0 | 11.1 | 0.0 | 5.0 | 0.0 | 5.3 |
| Decrease in energy | 0.0 | 0.0 | 18.2 | 11.1 | 0.0 | 15.0 | 11.1 | 5.3 |
| Dizziness/ light-headed | 14.3 | 0.0 | 0.0 | 11.1 | 5.9 | 5.0 | 5.6 | 5.3 |
| Flushing/heat sensation | 0.0 | 10.0 | 0.0 | 22.2 | 5.9 | 10.0 | 0.0 | 15.8 |
| Headache | 14.3 | 10.0 | 0.0 | 22.2 | 11.8 | 10.0 | 5.6 | 15.8 |
| Increase in alertness/energy | 71.4 | 10.0 | 18.2 | 11.1 | 35.3 | 15.0 | 38.9 | 10.5 |
| Itching | 0.0 | 60.0 | 0.0 | 55.6 | 35.3 | 25.0 | 0.0 | 57.9 |
| Muscle soreness/tightness | 85.7 | 20.0 | 45.5 | 33.3 | 47.1 | 40.0 | 61.1 | 26.3 |
| Pain > 2min after injection | 71.4 | 40.0 | 18.2 | 66.7 | 52.9 | 40.0 | 38.9 | 52.6 |
| Redness | 14.3 | 50.0 | 0.0 | 55.6 | 35.3 | 25.0 | 5.6 | 52.6 |
| Swelling | 14.3 | 0.0 | 0.0 | 22.2 | 5.9 | 10.0 | 5.6 | 10.5 |
| Tiredness/sleepy | 28.6 | 20.0 | 27.3 | 22.2 | 23.5 | 25.0 | 27.8 | 21.1 |
| Warmth | 28.6 | 30.0 | 18.2 | 11.1 | 29.4 | 15.0 | 22.2 | 21.1 |
